# Model-based meta-analysis to optimise *S. aureus*-targeted therapies for atopic dermatitis

**DOI:** 10.1101/2021.10.04.21264531

**Authors:** Takuya Miyano, Alan D Irvine, Reiko J Tanaka

## Abstract

**Background:** Several clinical trials of *Staphylococcus aureus* (*S. aureus*)-targeted therapies for atopic dermatitis (AD) have demonstrated conflicting results regarding whether they improve AD severity scores. This study performs a model-based meta-analysis to investigate possible causes of these conflicting results and suggests how to improve the efficacies of *S. aureus*-targeted therapies.

**Methods:** We developed a mathematical model that describes systems-level AD pathogenesis involving interactions between *S. aureus* and Coagulase Negative *Staphylococcus* (CoNS). The model was calibrated to reproduce time course data of *S. aureus* levels, EASI scores, and EASI-75 in response to dupilumab, *S. hominis* A9 (*Sh*A9) and flucloxacillin from published clinical trials. We simulated efficacies of hypothetical *S. aureus*-targeted therapies on virtual patients using the model.

**Results:** Our model simulation reproduced the clinically observed detrimental effects that application of *Sh*A9 and flucloxacillin had on AD severity and showed that these effects disappeared if the bactericidal activity against CoNS was removed. A hypothetical (modelled) eradication of *S. aureus* by 3.0 log_10_ CFU/cm^2^, without killing CoNS, achieved comparable EASI-75 to dupilumab. This efficacy was potentiated if dupilumab was administered in conjunction with *S. aureus* eradication (EASI-75 at week 16; *S. aureus* eradication: 66.7%, dupilumab 61.6% and combination: 87.8%). The improved efficacy was also seen for virtual dupilumab poor responders.

**Conclusion:** Our model simulation suggests that killing CoNS worsens AD severity and that *S. aureus*-specific eradication without killing CoNS could be effective for AD patients, including dupilumab poor responders. This study will contribute to design promising *S. aureus*-targeted therapy.

## 1. INTRODUCTION

Atopic dermatitis (AD), also called eczema, is the most common inflammatory skin disease^1^. The symptoms of AD involve relapsing pruritus and skin pain, impairing patients’ quality of life and work productivity^2^. The pathogenesis of AD is characterised by skin barrier damage, Th2-dominant inflammation and skin dysbiosis^3,4,5^. The skin dysbiosis in AD patients is frequently accompanied by *Staphylococcus aureus* (*S. aureus*) colonisation and decreased commensal bacteria in the skin^6^. *S. aureus* skin colonisation is found in 75%-90% of AD patients without clinical signs of superinfection, whereas it is found in only 0%-25% of healthy subjects^7-11^.

*S. aureus* levels on skin lesions correlate with AD severity^12,13^, and *S. aureus* has been considered as a promising target for AD treatment as it induces both skin barrier damage and inflammation by producing various virulence factors, such as phenol-soluble modulins (PSMs), staphylococcal enterotoxins and the toxic shock syndrome toxin-1^14,15^.

Some clinical trials of *S. aureus*-targeted therapies for AD^16^ have indeed demonstrated a reduction in *S. aureus*. However, they have shown conflicting efficacies as to whether they improve AD severity scores. For example, in several clinical trials, oral and topical antibiotics whose antibacterial spectrum covers *Staphylococci* were applied to eradicate *S. aureus* at least temporarily on AD skin lesions. But these interventions often failed to improve AD severity: A Cochrane review concluded that antibiotics may make no difference or only slight improvement in AD severity^17^. Oral flucloxacillin, one of the antibiotics, worsened AD severity compared to placebo despite a significant reduction of *S. aureus* levels on skin lesion^18^. Currently, the use of antibiotics is recommended for AD only in case of overt infection^19^.

As another *S. aureus*-targeted therapy, transplant of *S. hominis* A9 (*Sh*A9), a commensal strain of coagulase-negative staphylococci (CoNS) isolated from healthy human skin, has been tested^20^. The clinical study showed that *Sh*A9 transplant decreased the *S. aureus* levels on skin lesions and improved AD severity scores in the patients (*N*=21) whose skin was colonised with *S. aureus* that is sensitive to the bacteriocins secreted by *Sh*A9. However, *Sh*A9 transplant deteriorated AD severity scores in the patients (*N*=11) whose skin was colonised with *S. aureus* that is resistant to the bacteriocins secreted by *Sh*A9^20^. *Sh*A9 produces bacteriocins that have bactericidal activity against *S. aureus*^21^ and secretes autoinducing peptides (AIPs) that inhibit the accessory gene regulatory (agr) system, which regulates the expression of the virulence factors in *S. aureus*^22^.

Some therapeutics that do not target *S. aureus* directly can also reduce *S. aureus* levels. Dupilumab, an approved biologic for AD, is a monoclonal antibody that inhibits IL-4 and IL-13 signalling. These Th2 cytokines can facilitate *S. aureus* colonisation as they damage the skin barrier by inhibiting epidermal differentiation^23,24^and the skin barrier damage induces an increase in skin pH^25^ that promotes *S. aureus* growth^26^. In addition, inhibition of IL-4 and IL-13 by dupilumab can reduce *S. aureus* levels since IL-4 and IL-13 inhibit the synthesis of antimicrobial peptides (AMPs) against *S. aureus*^27^. Dupilumab has reduced *S. aureus* levels and improved AD severity scores in a clinical trial^12^.

Taken together, all of flucloxacillin, *Sh*A9 and dupilumab decreased *S. aureus* levels but showed conflicting efficacies as to whether these drugs improve AD severity scores. Understanding of underlying mechanism for the conflicting efficacies will help optimising *S. aureus*-targeted therapies for AD that are consistently effective.

To investigate the causes of the conflicting efficacies of *S. aureus*-targeted therapies, this study applies a quantitative systems pharmacology (QSP) approach. QSP is a framework to describe systems-level pathogenesis and drug effects by integrating data and knowledge into a mathematical model^28^. A QSP approach facilitates a model-based meta-analysis that integrates data from different clinical trials, as well as knowledge on pathogenesis and mechanism of action (MoA) of drugs, to inform rational drug development^29^. A QSP model-based meta-analysis is especially suitable for this study which aims to investigate underlying mechanisms for the conflicting efficacies of *S. aureus*-targeted therapies observed in different clinical studies.

We have recently applied a QSP model-based meta-analysis of multiple biologics for AD and identified IL-13 and IL-22 as potential drug targets for dupilumab poor responders^30^. However, the previous QSP model of biologics is not suitable for this study’s aim as it did not describe the mechanism of *S. aureus*-targeted therapies. This study presents a new QSP model of *S. aureus*-targeted therapies that describes the interactions between *S. aureus* and CoNS in AD pathogenesis by referring to clinical efficacy data of the three drugs described above (flucloxacillin, *Sh*A9 and dupilumab) to test the following two hypotheses.

The first hypothesis is that the bactericidal effects of *S. aureus*-targeted therapies on CoNS impair their efficacies on AD severity. Decrease in CoNS levels causes reduction in their AIP secretion, thereby upregulating agr expression. Upregulated agr expression promotes production of virulence factors in *S. aureus* that can worsen AD severity. While such a hypothesis has already been implied in several studies^20,31,32^, there has been no quantitative evaluation on the possible dynamic influences of killing CoNS on clinical efficacies to the best of our knowledge.

The second hypothesis is that *S. aureus*-targeted therapies are effective for dupilumab poor responders as they have a different MoA from dupilumab. The responder rates for dupilumab were 44%-69%^33,34^ for Eczema Area and Severity Index (EASI)-75 (75% reduction in the EASI score^35,36^), leaving a significant proportion of dupilumab poor responders. Therapeutic options for dupilumab poor responders are limited to increasing topical corticosteroids and adding additional systemic immunosuppressive agents. However, the dupilumab poor responders are often resistant to these treatments and require monitoring for adverse effects^37^, leaving unmet medical needs for dupilumab poor responders. We propose promising *S. aureus*-targeted therapies for AD patients, especially for dupilumab poor responders, by conducting model simulations on virtual patients.

## 2. METHODS

Our QSP model explicitly describes causal relationships between drugs, biological factors and an AD severity score using a graphical scheme and ordinary differential equations. The model was developed by 1) selecting drugs and biological factors to be modelled, 2) formulating drug effects and causal relationships between the biological factors and 3) optimising model parameters that define virtual patients. The developed model was used to simulate the clinical efficacies of hypothetical *S. aureus*-targeted therapies in virtual patients.

### 2.1. Selecting drugs and biological factors

We considered flucloxacillin, *Sh*A9 and dupilumab because they demonstrated a decrease of *S. aureus* levels in a placebo-controlled double-blinded clinical study where AD severity scores were reported (Table 1 and Supplementary Information (SI) Section 1).

**TABLE 1.**
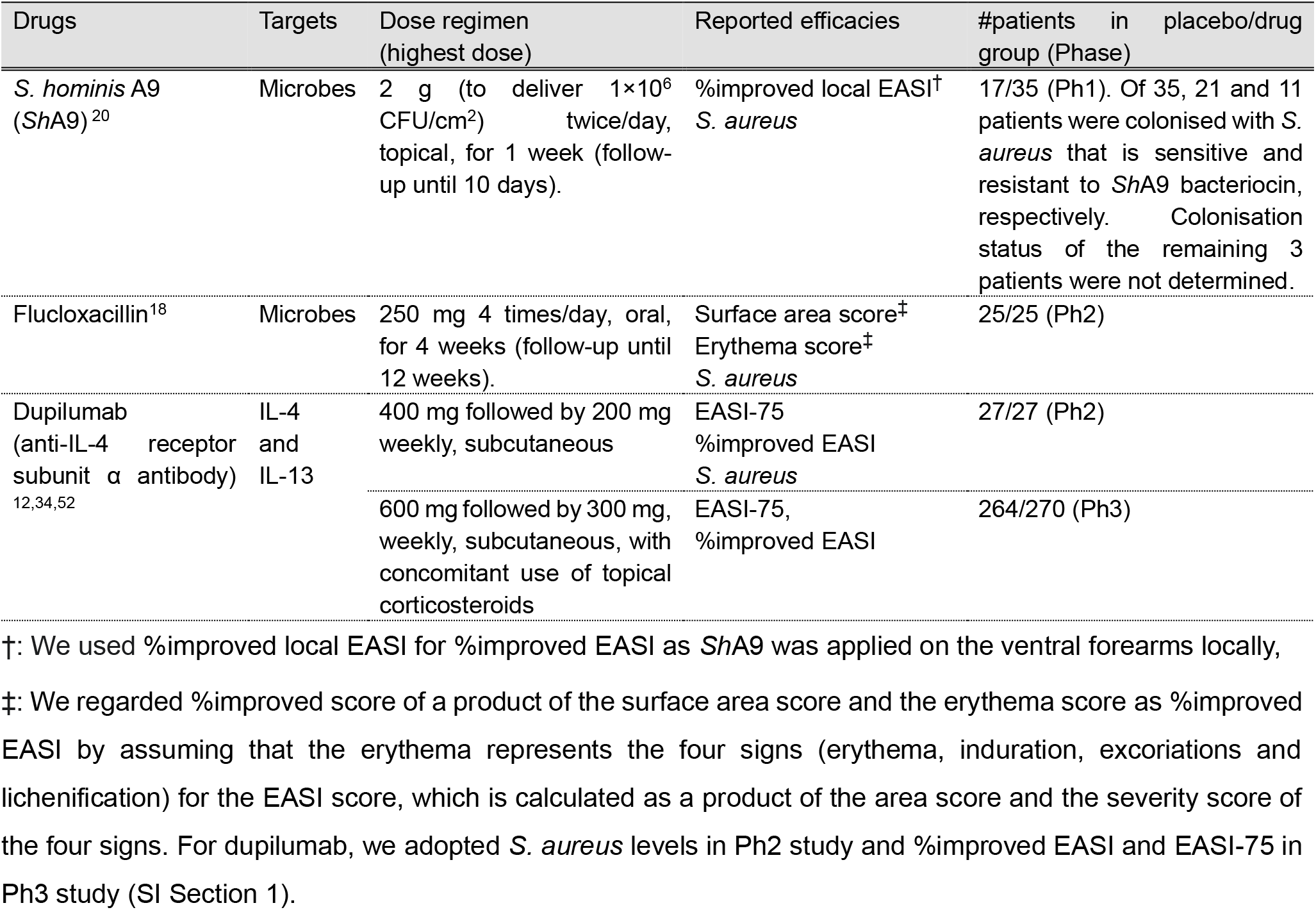
Drugs considered in this study

We selected six biological factors as model variables: colony density levels of *S. aureus* and CoNS and levels of agr expression, IL-4/IL-13 in the skin and skin barrier integrity and the EASI score. *S. aureus* and CoNS are the core factors in this study. “CoNS” does not include the *Sh*A9 strain applied in the *Sh*A9 treatment. “Agr expression*”* corresponds to the main mechanism for *S. aureus* to expression virulence factors in *S. aureus*^22^ that induce skin barrier damage and skin inflammation. The IL-4/IL-13 represents Th2-cytokines that are targeted by dupilumab. “Skin barrier integrity” is a critical factor in AD pathogenesis as in our previous models^30,38^. The EASI score represents an endpoint for AD severity. Some biological factors such as AMPs were not described as model variables but were considered implicitly as a rationale for the causal relationships (e.g., IL-4 and IL-13 increase *S. aureus* and CoNS via decreasing AMPs) to make the model simpler yet interpretable.

### 2.2. Formulating drug effects and causal relationships between biological factors

We developed a mathematical model consisting of six equations corresponding to the six biological factors with 26 parameters to simulate the efficacies of the three drugs (SI Section 3). The effects of flucloxacillin were modelled by increasing the killing rates of both *S. aureus* and CoNS as its antibacterial spectrum covers all Staphylococcus species. The effects of *Sh*A9 were modelled by increasing the killing rates of *S. aureus* and CoNS and the inhibitory strength against the agr expression because *Sh*A9 produces bacteriocins against both *S. aureus* and CoNS^21^ and AIPs that inhibit the agr expression^20^. The effects of dupilumab were modelled by decreasing effective concentrations of IL-4/IL-13 in the skin by 99%. The value of 99% was obtained from a calculation using the published data on IC_50_ and the mean concentration of drugs in the skin^39^ that was estimated from their concentration in the serum measured in clinical trials (SI section 3.2.3). The causal relationships between biological factors were described according to published experimental evidence based on human data (SI section 3.1). The model was implemented in Python 3.7.6 (Python Software Foundation).

### 2.3. Modelling virtual patients and optimising model parameters

We assumed that the model parameter values (e.g., the recovery rate of skin barrier via skin turnover, *k*_1_) vary between AD patients and that a set of 26 parameter values defines pathophysiological backgrounds of each virtual patient (TABLE S3). Each value of the *i*-th parameter, *k*_*i*_, is taken from a log-normal distribution^40^ whose probability function, *f*(*k*_*i*_), is defined by

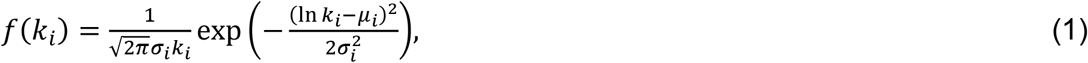

where *μ*_*i*_ and *σ*_*i*_ are the distribution parameters that represent the mean and the standard deviation of In *k*_*i*_, respectively.

We optimised the 52 distribution parameters (*μ*_*i*_ and *σ*_*i*_, *i*=1, …, 26) that define distributions of the 26 model parameters so that the model reproduces the reference data derived from published clinical studies (SI Section 4). The reference data consist of baseline levels of *S. aureus*, CoNS, IL-4/IL-13 and the EASI scores (TABLE S2) and time courses of *S. aureus* levels, the EASI scores and EASI-75 assessed in clinical trials of the selected drugs (FIGURE 1). The *S. aureus* levels, EASI scores and EASI-75 were normalised to compare the clinical efficacies of different clinical trials (SI Section 2). The agr expression and skin barrier integrity were regarded as latent state variables that have no reference data to be compared with simulated values. Simulated baseline levels were obtained by computing steady-state levels of biological factors (at 1000 weeks without drug treatment). All the simulations were conducted on 1000 virtual patients generated by randomly sampling each parameter value from the distribution in Eq. (1).

**FIGURE 1.**
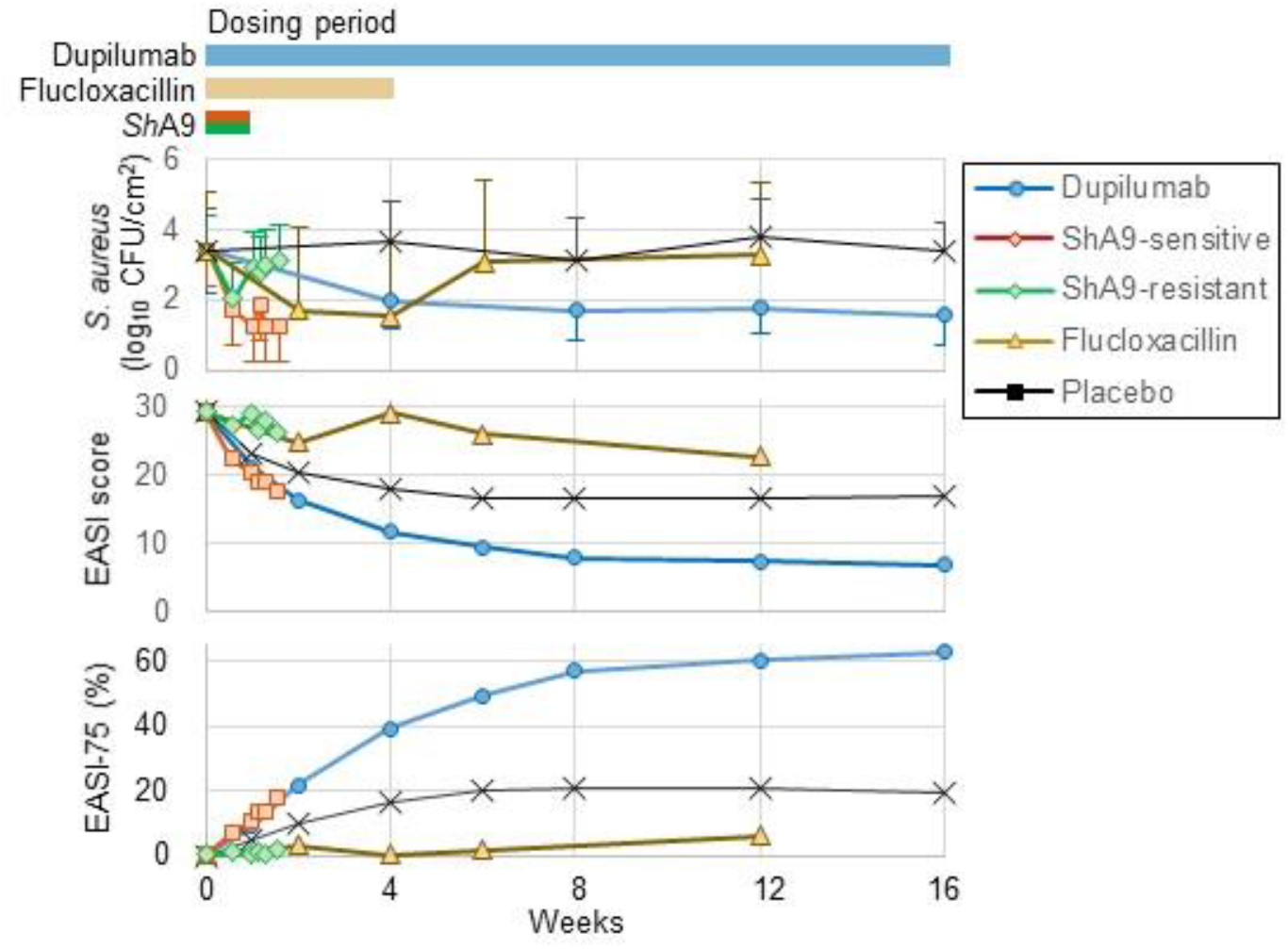
Three drugs (flucloxacillin, *Sh*A9 and dupilumab) reduced *S. aureus* levels but demonstrated conflicting clinical efficacies regarding EASI scores. *S. aureus* levels, the EASI score and EASI-75 were normalised using the reported data of each clinical trial (SI Section 2). For *Sh*A9, we evaluated the efficacies for the patients stratified by whether the colonised *S. aureus* is sensitive to *Sh*A9 bacteriocins (*Sh*A9-sensitive) or is resistant to *Sh*A9 bacteriocins (*Sh*A9-resistant). Horizontal bars on top represent the dosing periods in each clinical trial. Error bars: standard deviation.

### 2.4. Simulating efficacies of hypothetical *S. aureus*-targeted therapies

We simulated EASI-75 of hypothetical therapies with different strengths for killing of *S. aureus* and of CoNS and for inhibiting agr expression to explore optimal *S. aureus*-targeted therapies. Specifically, we examined the efficacies of hypothetical therapies that achieve a maximal reduction of *S. aureus* level from placebo (reduction of 3.0 log_10_ CFU/cm^2^ achieved in published clinical trials for *S. aureus*-targeted therapies^18, 20, 41, 42, 43, 44, 45^), the maximal level of CoNS (no bactericidal effects on CoNS, keeping the baseline level of CoNS), an example level of inhibition of the agr expression (we used 90% as we have no reliable evidence to estimate maximal inhibition rates of agr expression) and their combinations.

We also simulated EASI-75 of hypothetical therapies in virtual dupilumab poor responders, which were defined as the virtual patients who did not achieve the EASI-75 criterion at 16 weeks.

## 3. RESULTS

### 3.1. QSP model reproduced clinical efficacies of three drugs

We normalised *S. aureus* levels, the EASI scores and EASI-75 using the reported results in the clinical trials to compare efficacies of flucloxacillin, *Sh*A9 and dupilumab (FIGURE 1). The efficacies of *Sh*A9 were presented for two groups of patients stratified by the sensitivity of *S. aureus* to *Sh*A9 bacteriocins, as in the original clinical study^20^. Hereafter, *Sh*A9 applied to patients colonised with *S. aureus* that is sensitive to *Sh*A9 bacteriocins is referred as *Sh*A9-sensitive, and those with *S. aureus* that is resistant to *Sh*A9 bacteriocins is referred as *Sh*A9-resistant.

The normalised efficacies demonstrated that all the drugs decreased *S. aureus* levels and that *Sh*A9-sensitive and dupilumab improved the EASI scores and EASI-75, whereas *Sh*A9-resistant and flucloxacillin worsened the EASI scores and EASI-75. The results confirmed that the three drugs demonstrated conflicting efficacies regarding whether they improve AD severity scores while they all reduced *S. aureus* levels.

We revised our previously published QSP model of biolgics^30^ to include the MoA for the three drugs and the interactions between *S. aureus* and CoNS (FIGURE 2). The new QSP model of *S. aureus*-targeted therapies reproduced the baseline levels of the biological factors and the clinical efficacies of the drugs on *S. aureus* levels, the EASI scores and EASI-75 (FIGURE 3a, b). The root mean square errors of the mean and %CV of *S. aureus* levels, the EASI scores and EASI-75 between the simulated and reference data were 0.3 log_10_ CFU/cm^2^, 43%, 1.5 (out of 72 = the maximal EASI score) and 2.9%, respectively.

**FIGURE 2.**
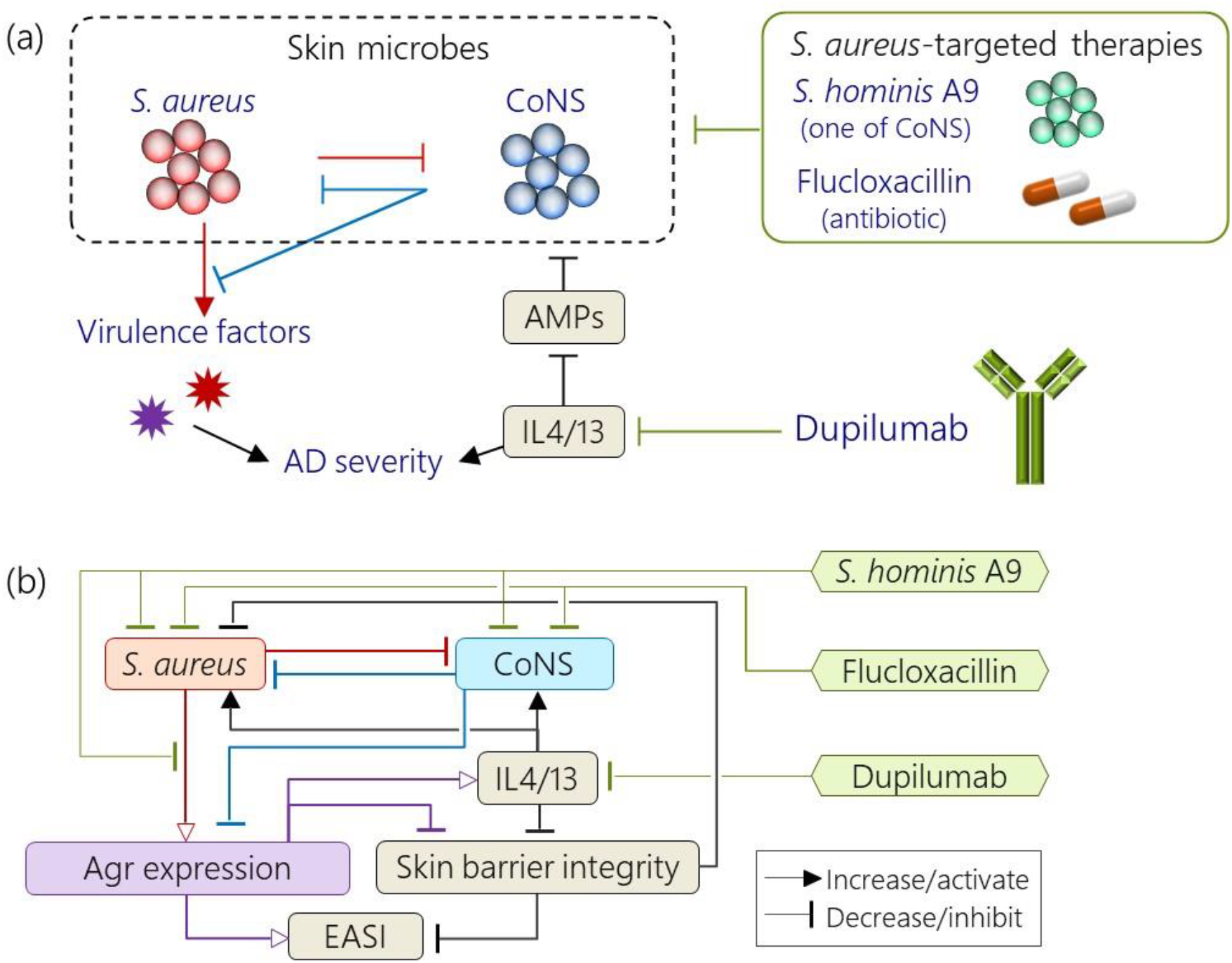
Overview of the QSP model that describes the interactions between *S. aureus* and CoNS in AD pathogenesis. (a) Schematic diagram. (b) Regulatory pathways of the QSP model. The model comprises of the EASI score (an efficacy endpoint), skin barrier integrity, agr expression, *S. aureus*, CoNS, IL-4/IL-13 and drugs (*Sh*A9, flucloxacillin, dupilumab). The regulatory pathways between biological factors are described according to published human data (SI Section 3).

**FIGURE 3.**
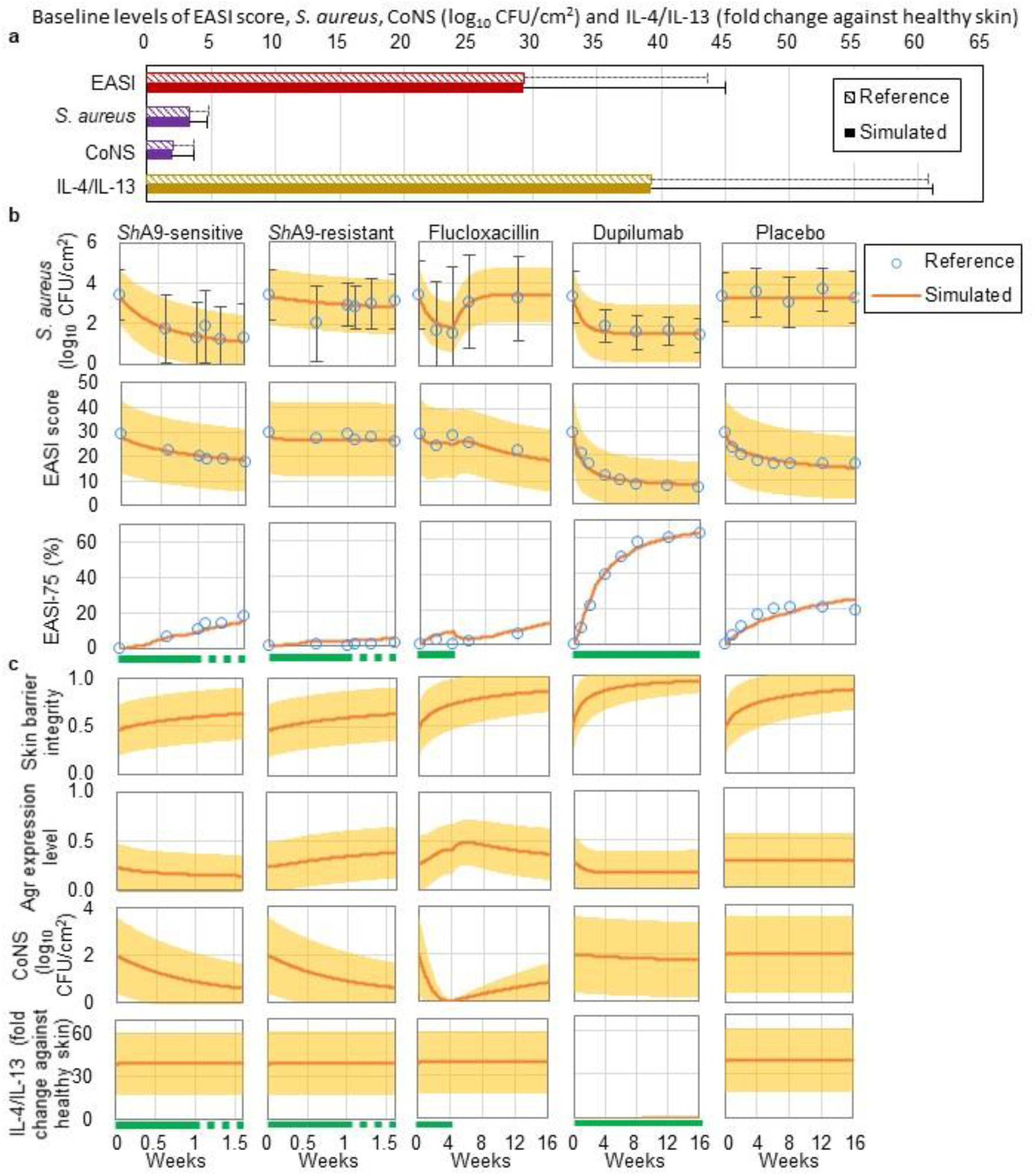
QSP model-based simulation reproduced the reference data. The distributions of the model parameters were optimised to minimize the difference between simulated and reference data (SI section 4). Simulation was conducted on 1000 virtual patients. **(a)** Comparison of baseline levels of biological factors between reference (striped bars) and simulated data (filled bars). Error bars: standard deviation. **(b)** Comparison of clinical efficacies of flucloxacillin, *Sh*A9 and dupilumab between reference (unfilled circles: mean. error bars: standard deviation) and simulated (lines: mean. shaded area: standard deviation .) data. **(c)** Simulated model variables that have no reference data (lines: mean. shaded area: standard deviation .). The IL-4/IL-13 levels in dupilumab reflect the 99% inhibition of IL-4/IL-13 by dupilumab. Green lines represent dosing periods. Effects of *Sh*A9 were applied in both dosing and follow-up periods in the simulation because the measured amounts of *Sh*A9 on the skin remained higher than baseline levels during the follow-up periods in the actual clinical trial^20^, while effects of flucloxacillin and dupilumab were applied only during dosing periods.

### 3.2. Detrimental effects of flucloxacillin and *Sh*A9 on EASI scores disappeared when their bactericidal activity against CoNS was hypothetically removed

Using the new QSP model, we tested the first hypothesis that the bactericidal effects on CoNS impair the efficacies of *S. aureus*-targeted therapies on AD severity.

Our model simulation demonstrated that flucloxacillin and *Sh*A9-resistant decreased CoNS while increasing the agr expression (FIGURE 3c), and that flucloxacillin and *Sh*A9 could achieve better EASI scores and EASI-75 than placebo if they had no bactericidal effects on CoNS (FIGURE 4). In addition, a sensitivity analysis of the model parameters for %improved EASI elucidated that lower rates of CoNS killing by flucloxacillin (*d*_fh_) and *Sh*A9 treatments (*d*_A9h_) result in higher %improved EASI (SI section 5). These results suggested that a decrease in CoNS increases the agr expression, thereby worsening the EASI scores.

**FIGURE 4.**
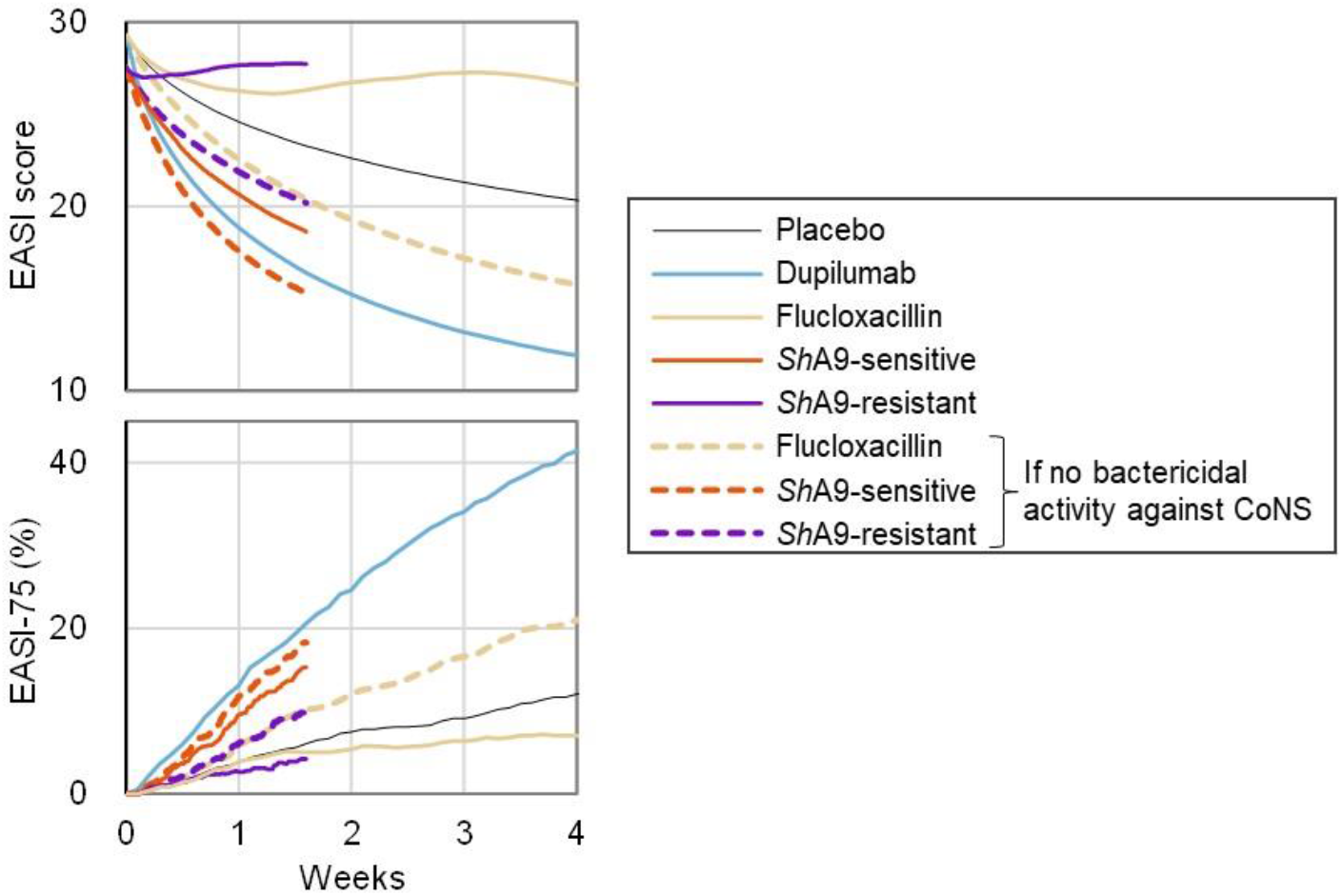
Detrimental effects of flucloxacillin and *Sh*A9 on EASI scores disappeared if their bactericidal activity against CoNS were hypothetically removed. The EASI scores and EASI-75 of flucloxacillin and *Sh*A9 (yellow, red and purple solid lines) were compared with hypothetical situation where flucloxacillin and *Sh*A9 have no bactericidal effects on CoNS (yellow, red and purple dashed lines). The efficacies of dupilumab (blue solid line), the effects of which were modelled by inhibiting IL-4/IL-13 by 99%, were shown as a reference. Simulation was conducted on 1000 virtual patients (The EASI scores: mean values. EASI-75: responder rates). Without bactericidal effects on CoNS, flucloxacillin and *Sh*A9 achieved better efficacies than placebo (black thin line) in our simulation. The simulation of efficacies of *Sh*A9 was stopped on day 10 because our model was calibrated to reproduce the reported efficacies of *Sh*A9 until day 10.

While CoNS levels were reduced to similar levels in both the *Sh*A9-sensitive and *Sh*A9-resistant groups, agr expression was reduced only in the *Sh*A9-sensitive group (FIGURE 3c). The agr expression decreased due to the stronger decrease of *S. aureus* levels by *Sh*A9-sensitive, compared to *Sh*A9-resistant, even though the decrease in CoNS resulted in a slight increase in the agr expression. These results suggest that the efficacies of *S. aureus*-targeted therapies are determined in some part by the balance of their bactericidal strengths against *S. aureus* vs. CoNS.

### 3.3. Hypothetical *S. aureus*-targeted therapies achieved better EASI-75 than dupilumab

The QSP model described antimicrobial effects of *S. aureus*-targeted therapies by three parameters: the rate of *S. aureus* killing, that of CoNS killing and the strength of agr expression inhibition (FIGURE 2). The antimicrobial effects result in a decrease of *S. aureus* level, that of CoNS level and an inhibition of agr expression level, respectively (FIGURE 5a). To explore which antimicrobial effects are responsible for improvement in AD severity, we conducted model simulations for hypothetical *S. aureus*-targeted therapies with different values of the three parameters.

**FIGURE 5.**
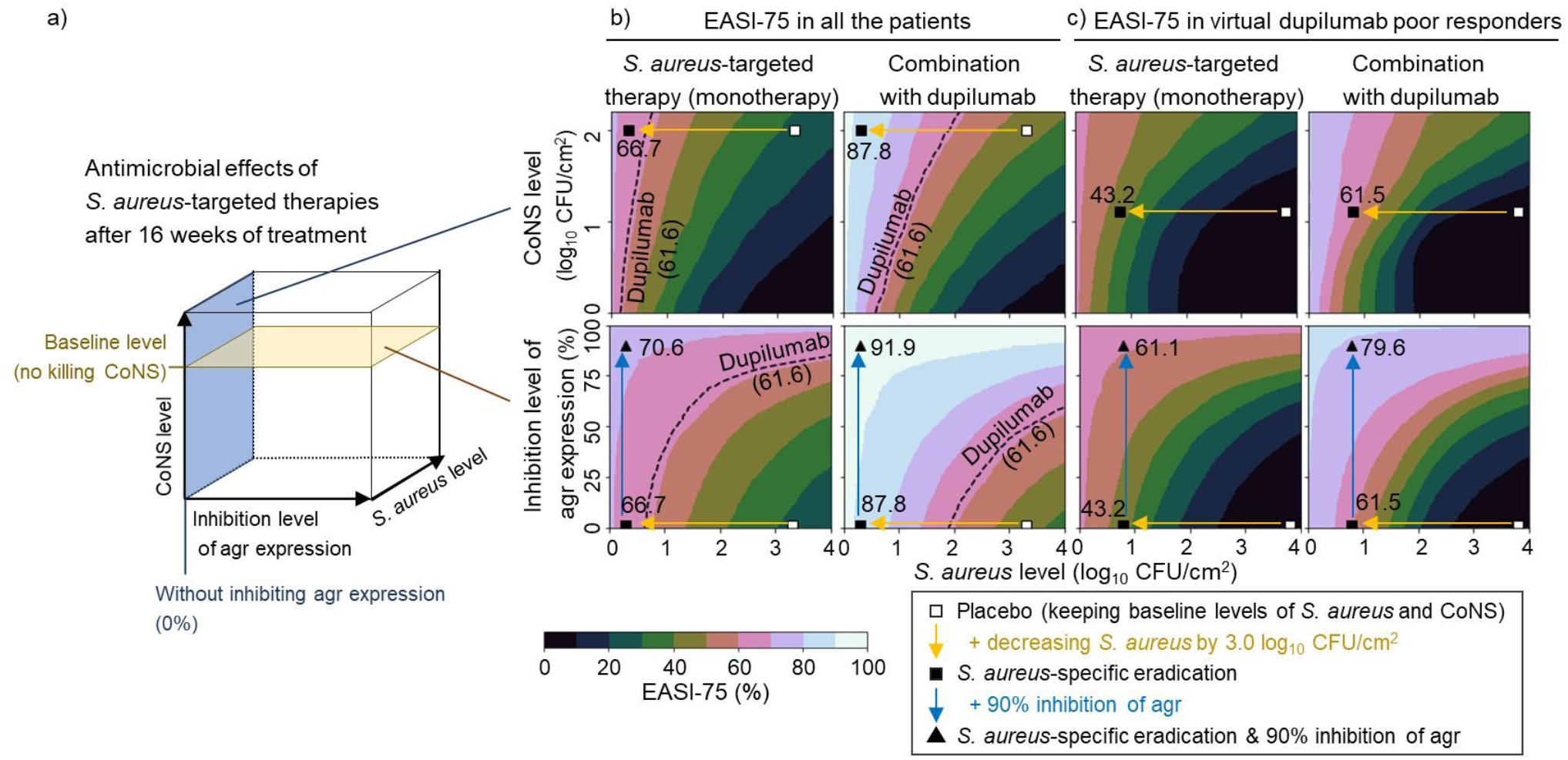
Hypothetical *S. aureus*-targeted therapies achieved better EASI-75 after 16 weeks of treatment than dupilumab in our model simulation. (a) Antimicrobial effects of hypothetical *S. aureus*-targeted therapies are represented by the level of *S. aureus*, that of CoNS and the inhibition level of agr expression after 16 weeks of treatment. Hypothetical *S. aureus*-targeted therapies were represented in our model by varying strengths of *S. aureus* killing, CoNS killing and inhibition of agr expression. (b and c) Antimicrobial effects of hypothetical *S. aureus*-targeted therapies evaluated by EASI-75 after 16 weeks of treatment for all virtual patients (b) and for virtual dupilumab poor responders (c). Lower *S. aureus* levels, higher CoNS levels and stronger inhibition of agr expression achieved a better EASI-75. The hypothetical *S. aureus*-specific eradication (yellow arrows) achieved comparable (b) or better (c) EASI-75 to dupilumab (dotted line in (b) and 0% in (c)), and its EASI-75 was potentiated (triangle) by adding 90% inhibition of agr expression (blue arrows). Their combination application with dupilumab achieved better EASI-75 than an application of either one. The effects of dupilumab were modelled by inhibiting IL-4/IL-13 by 99%. Simulation was conducted on 1000 virtual patients or 1000 virtual dupilumab poor responders (levels of *S. aureus* and CoNS and the inhibition level of agr expression: mean values. EASI-75: responder rates).

Our simulation results demonstrated that lower *S. aureus* levels, higher CoNS levels and stronger inhibition of agr expression resulted in higher EASI-75 after 16 weeks (FIGURE 5b left). The *S. aureus*-specific eradication (the maximal reduction of *S. aureus* level without killing CoNS, yellow arrows in FIGURE 5b) led to comparable EASI-75 to dupilumab (66.7% vs. dupilumab: 61.6%). The EASI-75 of the *S. aureus*-specific eradication was improved by adding 90% inhibition of the agr expression (70.6%, blue arrows in FIGURE 5b).

Simulations for a combinatorial application of dupilumab and hypothetical *S. aureus*-targeted therapies elucidated that it can achieve better EASI-75 than an application of either one (FIGURE 5b right). The *S. aureus*-specific eradication improved EASI-75 (87.8%) when it was combined with dupilumab, which was further improved (91.9%) by adding 90% inhibition of agr expression.

### 3.4. *S. aureus*-targeted therapies achieved significant response in virtual dupilumab poor responders

We also simulated EASI-75 of *S. aureus*-targeted therapies in dupilumab poor responders (FIGURE 5c). Similarly to the results shown above for all virtual patients (FIGURE 5b), lower *S. aureus* levels, higher CoNS levels and higher inhibition of agr expression showed a better EASI-75 in virtual dupilumab poor responders. The hypothetical *S. aureus*-targeted therapies achieved a significant EASI-75 in virtual dupilumab poor responders (*S. aureus*-specific eradication: 43.2% and that with 90% inhibition of agr expression: 61.1%), which were potentiated by simultaneous application of dupilumab (*S. aureus*-specific eradication: 61.5% and that with 90% inhibition of agr expression: 79.6%).

## 4. DISCUSSION

### 4.1. QSP model-based meta-analysis reveals mechanism of conflicting efficacies of *S. aureus*-targeted therapies

We developed a QSP model that describes the interactions between *S. aureus* and CoNS in AD pathogenesis (FIGURE 2) by integrating data and knowledge from published experiments using human samples (SI section 3). The model reproduced published data of clinical efficacy for flucloxacillin, *Sh*A9 and dupilumab (FIGURE 1) regarding the EASI scores, EASI-75 and *S. aureus* levels (FIGURE 3).

The QSP model simulation revealed that *S. aureus*-targeted therapies can worsen the EASI scores if they kill CoNS. The simulation showed that the application of *Sh*A9 and flucloxacillin had detrimental effects on AD severity, and those effects disappeared if their bactericidal activity against CoNS was hypothetically removed (FIGURE 4). The graphical scheme of the QSP model (FIGURE 2) can explain how a decrease in CoNS impairs the EASI scores. The decreased CoNS levels diminish secreted AIPs, thereby upregulating the agr expression. The upregulated agr expression promotes the production of virulence factors that damage the skin barrier (e.g., by PSMα and enterotoxins) and induce inflammation (e.g., by wall teichoic acid to activate dendritic cells), which can worsen AD severity. These results and interpretation indicate an importance of bactericidal specificity on *S. aureus* in *S. aureus*-targeted therapies.

### 4.2. Model simulation quantifies relationships between profiles of antibacterial effects and responder rates

The QSP model simulation also elucidated relationships between antibacterial effects of *S. aureus*-targeted therapies (decreases in the *S. aureus* and CoNS levels and in the inhibition level of agr expression) and their EASI-75 responder rates (FIGURE 5b left). In addition, our simulation suggested that the efficacy of *S. aureus*-targeted therapies can be potentiated by concomitant use of dupilumab (FIGURE 5b right).

Theoretically, *S. aureus*-targeted therapies will achieve the best efficacy if they could eradicate *S. aureus* completely. However, some *S. aureus* may remain on population average after *S. aureus*-targeted therapies, presumably due to resistance against antibiotics and bacteriocins^46^. Hence, it is crucial to inhibit agr expression by keeping the AIPs produced by CoNS, in addition to killing *S. aureus*, to minimise the agr-dependent virulence effects of *S. aureus*.

The hypothetical *S. aureus*-specific eradication (the maximal reduction of *S. aureus* level without killing CoNS), especially in combination with dupilumab, showed higher responder rates than dupilumab (Simulated EASI-75 at week 16: placebo 26.6%, dupilumab 61.6%, *S. aureus*-specific eradication 66.7% and combination 87.8%. FIGURE 5b right). Recently, JAK inhibitors have demonstrated promising efficacies in AD patients; abrocitinib showed a comparable response to dupilumab (EASI-75 at week 16; abrocitinib 71.0% vs. dupilumab 65.5%, not significant)^47^, and upadacitinib showed the highest responder rate among Ph3 trials of JAK inhibitors (EASI-75 at week 16. upadacitinib 77.1% vs. placebo 26.4%)^48^. Our simulation implies that *S. aureus*-specific eradication combined with dupilumab may achieve higher responder rates than JAK inhibitors.

### 4.3. *S. aureus*-specific eradication is potentially effective for dupilumab poor responders

This study also suggested the effectiveness of *S. aureus*-targeted therapies for dupilumab poor responders. The simulation for virtual dupilumab poor responders showed that *S. aureus*-specific eradication achieved 43.2% EASI-75 (FIGURE 5c left), which is much higher than EASI-75 achieved (up to 33.8%) when we simulated inhibition of all the cytokines considered in the previous QSP model of biologics^30^. These results imply that *S. aureus*, rather than cytokines, is potentially a promising therapeutic target for dupilumab poor responders.

The model simulation also demonstrated that the efficacy of *S. aureus*-targeted therapies is potentiated by its concomitant use with dupilumab in dupilumab poor responders (FIGURE 5c right). The results suggest that IL-4/IL-13 signalling contributes to the pathogenesis even for dupilumab poor responders and thus needs to be inhibited. Targeting both *S. aureus* and IL-4/IL-13 could be a promising therapeutic approach for AD patients.

### 4.4. Limitation of the QSP model simulation

This study aimed to interpret clinical data of *S. aureus*-targeted therapies obtained under different study conditions using a model-based meta-analysis. We assumed their efficacies are comparable across clinical trials after normalisation, although the study conditions (e.g., topical and systemic therapies) may influence the reported efficacies. The accuracy of the simulated efficacies of the hypothetical *S. aureus*-targeted therapies needs to be verified by future clinical trials^49^.

Our model assumed that CoNS has no detrimental effects on the skin barrier and inflammation. However, recent studies have suggested that *S. epidermidis*, one of CoNS, also has detrimental effects on the skin barrier^13^. The detrimental effects of *S. epidermidis* may explain the worsened EASI scores in *Sh*A9 as it increased the proportion of *S. epidermidis* among microbiome in the AD skin lesion^20^. Explicit modelling of different CoNS strains may deliver further insights into the roles of CoNS in AD pathogenesis, although our model assumed the detrimental effects of *S. epidermidis* are negligible compared to those of *S. aureus* because *S. aureus* has a higher correlation with AD severity scores than *S. epidermidis*^6,50^.

### 4.5. *Prospect for S. aureus*-targeted therapies

The results of this study supported the widely accepted idea for *S. aureus* being a promising drug target for AD and suggested the importance of considering antibacterial activities against both *S. aureus* and CoNS when developing *S. aureus*-targeted therapies. How much *S. aureus* killing is required to achieve a certain efficacy for a therapy would depend on how strong the therapy kills CoNS and inhibits agr expression.

This study presents an example of how QSP model can contribute to model-informed drug development^51^ for precision medicine. For example, our simulation results will contribute to the design of *S. aureus*-targeted therapies because the simulated relationship between EASI-75 responder rates and antibacterial effects (i.e., decreases in the *S. aureus* and CoNS levels and inhibition of agr expression) can be used as a guide to set a target profile of the antibacterial effects to achieve a desirable efficacy (e.g., better EASI-75 than dupilumab). Our simulation results also encourage combinatorial use of *S. aureus*-targeted therapies and cytokine-targeted therapies such as biologics and JAK inhibitors for AD.

The code of the QSP model is available at https://github.com/Tanaka-Group/AD_QSP_model.

## Data Availability

The code of the QSP model is available at https://github.com/Tanaka-Group/AD_QSP_model.

## Acknowledgments

We thank Dr. Elisa Domínguez-Hüttinger for her insightful comments on our manuscript.

## References

1. Deckers IA, McLean S, Linssen S, Mommers M, van Schayck CP, Sheikh A. Investigating international time trends in the incidence and prevalence of atopic eczema 1990-2010: a systematic review of epidemiological studies. PLoS One. 2012;7:e39803

2. Simpson EL, Bieber T, Eckert L, Wu R, Ardeleanu M, Graham NM, Pirozzi G, Mastey V. Patient burden of moderate to severe atopic dermatitis (AD): Insights from a phase 2b clinical trial of dupilumab in adults. J Am Acad Dermatol. 2016;74:491–8

3. Czarnowicki T, He H, Krueger JG, Guttman-Yassky E. Atopic dermatitis endotypes and implications for targeted therapeutics. J Allergy Clin Immunol. 2019;143:1–11

4. Langan SM, Irvine AD, Weidinger S. Atopic dermatitis. Lancet. 2020;396:345–360.

5. Weidinger S, Beck LA, Bieber T, Kabashima K, Irvine AD. Atopic dermatitis. Nat Rev Dis Primers. 2018;4:1

6. Ederveen THA, Smits JPH, Hajo K, et al. A generic workflow for Single Locus Sequence Typing (SLST) design and subspecies characterization of microbiota. Sci Rep. 2019;9(1):19834.

7. Breuer K, HAussler S, Kapp A, Werfel T. Staphylococcus aureus: colonizing features and influence of an antibacterial treatment in adults with atopic dermatitis. Br J Dermatol. 2002;147(1):55–61.

8. Gong JQ, Lin L, Lin T, et al. Skin colonization by Staphylococcus aureus in patients with eczema and atopic dermatitis and relevant combined topical therapy: a double-blind multicentre randomized controlled trial. Br J Dermatol. 2006;155(4):680–687.

9. Park HY, Kim CR, Huh IS, et al. Staphylococcus aureus Colonization in Acute and Chronic Skin Lesions of Patients with Atopic Dermatitis. Ann Dermatol. 2013;25(4):410–416.

10. Higaki S, Morohashi M, Yamagishi T, Hasegawa Y. Comparative study of staphylococci from the skin of atopic dermatitis patients and from healthy subjects. Int J Dermatol. 1999;38(4):265–269.

11. Nath S, Kumari N, Bandyopadhyay D, et al. Dysbiotic Lesional Microbiome With Filaggrin Missense Variants Associate With Atopic Dermatitis in India. Front Cell Infect Microbiol. 2020;10:570423.

12. Callewaert C, Nakatsuji T, Knight R, et al. IL-4Rα Blockade by Dupilumab Decreases Staphylococcus aureus Colonization and Increases Microbial Diversity in Atopic Dermatitis. J Invest Dermatol. 2020;140(1):191–202.e7.

13. Cau L, Williams MR, Butcher AM, et al. Staphylococcus epidermidis protease EcpA can be a deleterious component of the skin microbiome in atopic dermatitis. J Allergy Clin Immunol. 2021;147(3):955–966.e16.

14. Syed AK, Reed TJ, Clark KL, Boles BR, Kahlenberg JM. Staphlyococcus aureus phenol-soluble modulins stimulate the release of proinflammatory cytokines from keratinocytes and are required for induction of skin inflammation. Infect Immun. 2015;83(9):3428–3437.

15. Geoghegan JA, Irvine AD, Foster TJ. Staphylococcus aureus and Atopic Dermatitis: A Complex and Evolving Relationship. Trends Microbiol. 2018;26(6):484–497.

16. Tham EH, Koh E, Common JEA, Hwang IY. Biotherapeutic Approaches in Atopic Dermatitis. Biotechnol J. 2020;15(10):e1900322.

17. George SM, Karanovic S, Harrison DA, et al. Interventions to reduce Staphylococcus aureus in the management of eczema. Cochrane Database Syst Rev. 2019;2019(10):CD003871.

18. Ewing CI, Ashcroft C, Gibbs AC, Jones GA, Connor PJ, David TJ. Flucloxacillin in the treatment of atopic dermatitis. Br J Dermatol. 1998;138(6):1022–1029.

19. LePoidevin LM, Lee DE, Shi VY. A comparison of international management guidelines for atopic dermatitis. Pediatr Dermatol. 2019;36(1):36–65.

20. Nakatsuji T, Hata TR, Tong Y, et al. Development of a human skin commensal microbe for bacteriotherapy of atopic dermatitis and use in a phase 1 randomized clinical trial. Nat Med. 2021;27(4):700–709.

21. Nakatsuji T, Chen TH, Narala S, et al. Antimicrobials from human skin commensal bacteria protect against Staphylococcus aureus and are deficient in atopic dermatitis. Sci Transl Med. 2017;9(378):eaah4680.

22. Williams MR, Costa SK, Zaramela LS, et al. Quorum sensing between bacterial species on the skin protects against epidermal injury in atopic dermatitis. Sci Transl Med. 2019;11(490):eaat8329.

23. Seltmann J, Roesner LM, von Hesler FW, Wittmann M, Werfel T. IL-33 impacts on the skin barrier by downregulating the expression of filaggrin. J Allergy Clin Immunol. 2015;135(6):1659–61.e4

24. Howell MD, Kim BE, Gao P, et al. Cytokine modulation of atopic dermatitis filaggrin skin expression. J Allergy Clin Immunol. 2009;124(3 Suppl 2):R7–R12

25. Elias PM. The how, why and clinical importance of stratum corneum acidification. Exp Dermatol. 2017;26(11):999–1003.

26. Lambers H, Piessens S, Bloem A, Pronk H, Finkel P. Natural skin surface pH is on average below 5, which is beneficial for its resident flora. Int J Cosmet Sci. 2006;28(5):359–370.

27. Howell MD, Boguniewicz M, Pastore S, et al. Mechanism of HBD-3 deficiency in atopic dermatitis. Clin Immunol. 2006;121(3):332–338

28. Schoeberl B. Quantitative Systems Pharmacology models as a key to translational medicine. Curr Opin Syst Biol 2019;16:25–31

29. Gibbs JP, Menon R, Kasichayanula S. Bedside to Bench: Integrating Quantitative Clinical Pharmacology and Reverse Translation to Optimize Drug Development. Clin Pharmacol Ther. 2018;103(2):196–198

30. Miyano T, Irvine AD, Tanaka RJ. A mathematical model to identify optimal combinations of drug targets for dupilumab poor responders in atopic dermatitis. Allergy. 2021. doi:10.1111/all.14870

31. Clowry J, Irvine AD, McLoughlin RM. Next-generation anti-Staphylococcus aureus vaccines: A potential new therapeutic option for atopic dermatitis?. J Allergy Clin Immunol. 2019;143(1):78–81.

32. Katsuyama M, Kobayashi Y, Ichikawa H, Mizuno A, Miyachi Y, Matsunaga K, Kawashima M. A novel method to control the balance of skin microflora Part 2. A study to assess the effect of a cream containing farnesol and xylitol on atopic dry skin. J Dermatol Sci. 2005 Jun;38(3):207–13.

33. Simpson EL, Bieber T, Guttman-Yassky E, et al. Two Phase 3 Trials of Dupilumab versus Placebo in Atopic Dermatitis. N Engl J Med. 2016;375:2335–2348

34. Blauvelt A, de Bruin-Weller M, Gooderham M, et al. Long-term management of moderate-to-severe atopic dermatitis with dupilumab and concomitant topical corticosteroids (LIBERTY AD CHRONOS): a 1-year, randomised, double-blinded, placebo-controlled, phase 3 trial. Lancet. 2017;389:2287–2303

35. Hanifin JM, Thurston M, Omoto M, Cherill R, Tofte SJ, Graeber M. The eczema area and severity index (EASI): assessment of reliability in atopic dermatitis. EASI Evaluator Group. Exp Dermatol. 2001;10:11–18

36. Schram ME, Spuls PI, Leeflang MM, Lindeboom R, Bos JD, Schmitt J. EASI, (objective) SCORAD and POEM for atopic eczema: responsiveness and minimal clinically important difference. Allergy. 2012;67:99–106

37. Hendricks AJ, Lio PA, Shi VY. Management Recommendations for Dupilumab Partial and Non-durable Responders in Atopic Dermatitis. Am J Clin Dermatol. 2019;20:565–569

38. Domínguez-Hüttinger E, Christodoulides P, Miyauchi K, et al. Mathematical modeling of atopic dermatitis reveals “double-switch” mechanisms underlying 4 common disease phenotypes. J Allergy Clin Immunol. 2017;139:1861–1872.e7

39. Vazquez ML, Kaila N, Strohbach JW, et al. Identification of N-{cis-3-[Methyl(7H-pyrrolo[2,3-d]pyrimidin-4-yl)amino]cyclobutyl}propane-1-sulfonamide (PF-04965842): A Selective JAK1 Clinical Candidate for the Treatment of Autoimmune Diseases. J Med Chem. 2018;61:1130–1152

40. Limpert E, Stahel WA, Abbt M. Log-normal distributions across the sciences: keys and clues: on the charms of statistics, and how mechanical models resembling gambling machines offer a link to a handy way to characterize log-normal distributions, which can provide deeper insight into variability and probability-normal or log-normal: that is the question. BioScience. 2001;51:341–352

41. Boguniewicz M, Sampson H, Leung SB, Harbeck R, Leung DY. Effects of cefuroxime axetil on Staphylococcus aureus colonization and superantigen production in atopic dermatitis. J Allergy Clin Immunol. 2001;108(4):651–652.

42. Hung SH, Lin YT, Chu CY, et al. Staphylococcus colonization in atopic dermatitis treated with fluticasone or tacrolimus with or without antibiotics. Ann Allergy Asthma Immunol. 2007;98(1):51–56.

43. Leyden JJ, Kligman AM. The case for steroid--antibiotic combinations. Br J Dermatol. 1977;96(2):179–187.

44. Korting HC, Zienicke H, Braun-Falco O, et al. Modern topical glucocorticoids and anti-infectives for superinfected atopic eczema: do prednicarbate and didecyldimethylammoniumchloride form a rational combination?. Infection. 1994;22(6):390–394.

45. Breneman DL, Hanifin JM, Berge CA, Keswick BH, Neumann PB. The effect of antibacterial soap with 1.5% triclocarban on Staphylococcus aureus in patients with atopic dermatitis. Cutis. 2000;66(4):296–300.

46. Harkins CP, Holden MTG, Irvine AD. Antimicrobial resistance in atopic dermatitis: Need for an urgent rethink. Ann Allergy Asthma Immunol. 2019;122(3):236–240.

47. Bieber T, Simpson EL, Silverberg JI, et al. Abrocitinib versus Placebo or Dupilumab for Atopic Dermatitis. N Engl J Med. 2021;384(12):1101–1112.

48. Reich K, Teixeira HD, de Bruin-Weller M, et al. Safety and efficacy of upadacitinib in combination with topical corticosteroids in adolescents and adults with moderate-to-severe atopic dermatitis (AD Up): results from a randomised, double-blind, placebo-controlled, phase 3 trial. Lancet. 2021;397(10290):2169–2181.

49. Cucurull-Sanchez L, Chappell MJ, Chelliah V, et al. Best Practices to Maximize the Use and Reuse of Quantitative and Systems Pharmacology Models: Recommendations From the United Kingdom Quantitative and Systems Pharmacology Network. CPT Pharmacometrics Syst Pharmacol. 2019;8(5):259–272.

50. Byrd AL, Deming C, Cassidy SKB, et al. Staphylococcus aureus and Staphylococcus epidermidis strain diversity underlying pediatric atopic dermatitis. Sci Transl Med. 2017;9(397):eaal4651.

51. EFPIA MID3 Workgroup, Marshall SF, Burghaus R, et al. Good Practices in Model-Informed Drug Discovery and Development: Practice, Application, and Documentation. CPT Pharmacometrics Syst Pharmacol. 2016;5:93–122

52. Guttman-Yassky E, Bissonnette R, Ungar B, et al. Dupilumab progressively improves systemic and cutaneous abnormalities in patients with atopic dermatitis. J Allergy Clin Immunol. 2019;143(1):155–172.

