## Supplementary Information for "Model-based meta-analysis to optimise *S. aureus*-targeted therapies for atopic dermatitis"

### 1. Selection of clinical studies for development of the QSP model

We used pre-defined inclusion and exclusion criteria (FIGURE S1) to select clinical studies to be referenced in development of the QSP model. Firstly, we identified 24 clinical trials that reported both *S. aureus* levels and AD severity scores in a placebo-controlled study. We then excluded 18 clinical trials as the drugs have unclear MoA, they failed to decrease *S. aureus* levels compared to placebo, or they evaluated only a small number (<10) of patients. TABLE S1 lists the drugs excluded in this study. As for antibiotics/antiseptics, 23 clinical trials were investigated in Cochrane review[1], from which we included only one study[2] with flucloxacillin which met our inclusion criteria.

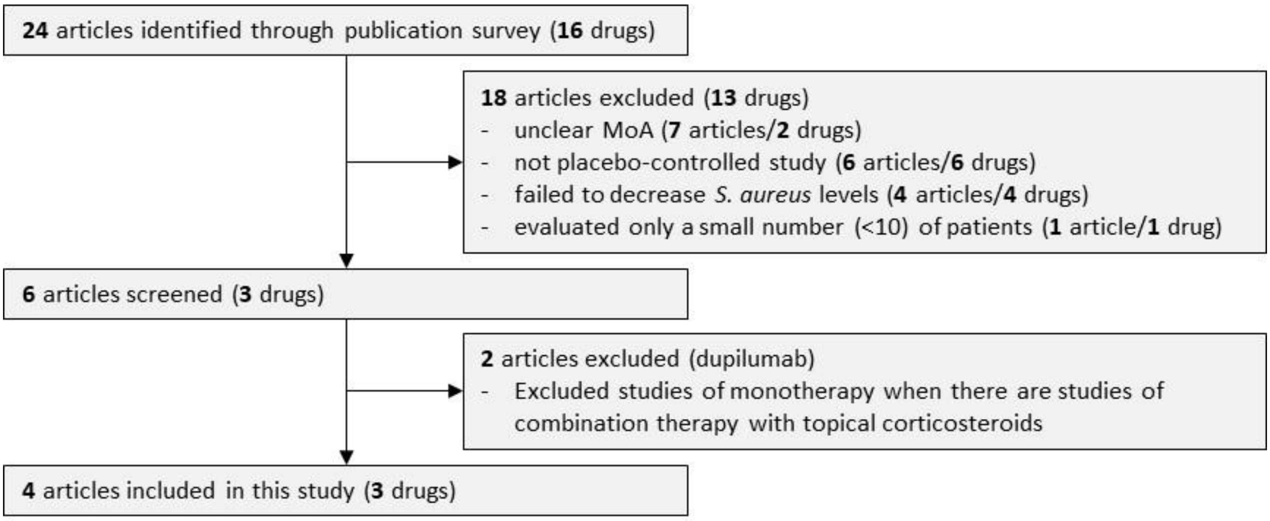


**FIGURE S1** Clinical studies selection process

**TABLE S1** Drugs excluded in this study (except for antibiotics/antiseptics)

| Drugs | MoA | Clinical efficacies (compared to placebo) | Reasons for exclusion |
|---|---|---|---|
| Bleach bath (hypochlorite 0.005%)[3] | Unclear (inhibiting NF-κB?) | Decreased *S. aureus* levels and improved EASI score | Unclear MoA; hypochlorite 0.005% inhibited NF-κB signaling in human keratinocytes, but was not antimicrobial against *S. aureus*[4,5]. |
| *Vitreoscilla filiformis* lysate[6] | Unclear (anti-inflammatory?) | Decreased *S. aureus* levels and improved SCORAD | Unclear MoA: target molecules are unknown |
| Staphefekt (Bacteriophage lysin)[7] | Killing *S. aureus* | Failed to decrease *S. aureus* levels and EASI score compared with placebo | Failed to decrease *S. aureus* levels compared with placebo control |
| *Roseomonas mucosa*[8] | Producing sphingolipid | Not a placebo-controlled study | Not a placebo-controlled study |
| Autologous CoNS[9] | Killing *S. aureus* via bacteriocins | Decreased *S. aureus* levels and improved EASI score | The number of subjects (5-6 subjects/arm) was too small |
| SRD441[10] (protease inhibitor) | Inhibiting Staphylococcal-derived aureolysin and matrix metalloproteinases | Slightly improved SCORAD without statistical significance. *S. aureus* levels were not reported | Not reported *S. aureus* levels |

As for dupilumab, we included the data of *S. aureus* levels in Ph2 study[11] and %improved EASI and EASI-75 in Ph3 study[12]. Detailed rationale for this choice is as follows.

1) %improved EASI and EASI-75 were reported in both Ph2 and Ph3 studies. However, *S. aureus* levels were reported only in Ph2 study[11].
2) The measured %improved EASI of placebo treatment in the Ph2 study[11] is not deemed to be reliable because the relationship between measured %improved EASI and EASI-75 at week 16 deviates from those in other clinical trials (FIGURE S2 left).
3) The estimated %improved EASI by mixed-effect model repeated measure (MMRM), which is a popular method to handling missing data (e.g., due to drop-out of patients during a clinical study)[13], in the Ph2 study[11] is deemed to be reliable because the relationship between the estimated %improved EASI and EASI-75 at week 16 in the Ph2 study[11] is consistent with those in other clinical trials (FIGURE S2 left). However, the estimated value is not time-course data (reported week 16 only) and cannot be used for our model fitting.
4) We decided to use a Ph3 study[12] that reported time-course data of %improved EASI and EASI-75, because (a) the %improved EASI at week 16 in the Ph3 study[12] is comparable to the estimated %improved EASI by MMRM at week 16 in the Ph2 study[11] (FIGURE S2 right, open and closed circles) and (b) time-course data of %improved EASI of dupilumab treatment in Ph2 were comparable to those in Ph3 (FIGURE S2 right, blue crosses and filled circles) suggesting that time-course data of %improved EASI of placebo treatment in Ph2, if they were estimated by MMRM, are comparable to those in Ph3**.**
5) Among several Ph3 studies for dupilumab[12, 14], we selected a Ph3 study which used a combination therapy with topical corticosteroids[12], as it is more reflective of the likely clinical use than monotherapy.

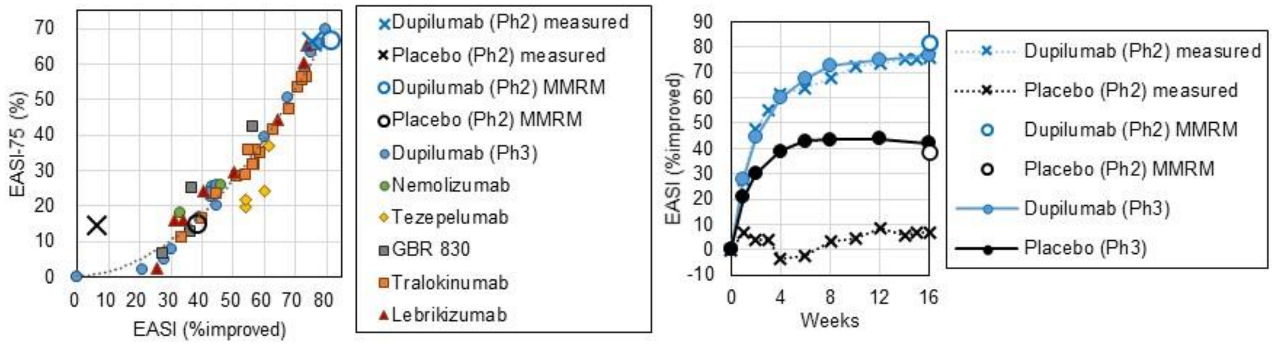


**FIGURE S2** %improved EASI reported in different clinical trials. (left) The relationship between %improved EASI and EASI-75. The placebo data measured at week 16 in a dupilumab Ph2 study[11] (a black cross) deviates from the data from other clinical trials (all available time points of both drug- and placebo-treated groups in dupilumab Ph3[12], nemolizumab[15], tezepelumab[16], GBR 830[17], lebrikizumab[18] and tralokinumab[19] studies) and the relationship between the %improved EASI estimated by mixed-effect model repeated measure (MMRM) and EASI-75 measured at week 16 in a dupilumab Ph2 study[11] (a blue open circle for dupilumab-treated and a black open circle for placebo-treated groups). (right) %improved EASI in dupilumab Ph2 and Ph3 studies. The estimated %improved EASI by MMRM at week 16 in Ph2[11] (open circles) is comparable to %improved EASI in Ph3[12] (filled circles) for both dupilumab- and placebo-treated groups.

### 2. Data processing

We used clinical efficacies (*S. aureus* levels, %improved EASI, EASI scores and EASI-75) of flucloxacillin, *S. hominis* A9 (*Sh*A9) and dupilumab as the reference data. The clinical efficacies were normalised to compare data from different clinical trials.

#### 2.1. Normalisation of *S. aureus* levels

Time courses of *S. aureus* levels were reported in clinical trials of flucloxacillin[2], *Sh*A9[20] and dupilumab[11]. We described the normalised *S. aureus* level, $a_j(t)$, for the *j*-th drug at time *t* by

$$a_j(t) = (\Delta a_j^*(t) - \Delta a_{\mathrm{P}_j}^*(t)) + \Delta a_{\mathrm{P_d}}^*(t) + a_{\mathrm{ShA9}}^*(0), \quad \text{(S1)}$$

where the first term corresponds to the net effects of the drug defined by the difference of the change in *S. aureus* levels ($\log_{10}$ scale) at time *t* from baseline, between the *j*-th drug ($\Delta a_j^*(t)$) and the corresponding placebo groups ($\Delta a_{\mathrm{P}_j}^*(t)$). This term adjusts for different placebo effects across clinical studies that may differ in the study participants' background, concomitant drugs and the study sites[21]. The remaining two terms describe the change in *S. aureus* levels at time *t* from baseline, $\Delta a_{\mathrm{P_d}}^*(t)$, in the placebo group in the dupilumab clinical trial that evaluated efficacies for the longest period among the trials evaluated in this study and the baseline level of *S. aureus*, $a_{\mathrm{ShA9}}^*(0)$, in the *Sh*A9 clinical trial that is the only one reporting levels of both *S. aureus* and CoNS among the trials evaluated in this study.

#### 2.2. Conversion of reported AD severity scores to %improved EASI

%improved EASI for flucloxacillin and *Sh*A9 were estimated as follows.

For flucloxacillin, we substituted %improved EASI by the %improved score of a product of the area score and the severity score of erythema[2] (the only disease sign evaluated in that study) by assuming that the erythema represents the four disease signs (erythema, induration, excoriations and lichenification) for EASI score, which is calculated as a product of the area score and the severity score of the four signs.

For *Sh*A9, we substituted %improved EASI by the %improved local EASI of the ventral arms as *Sh*A9 was applied on the ventral forearms locally[20].

#### 2.3. Normalisation of %improved EASI, EASI score and EASI-75

%improved EASI, EASI score and EASI-75 were normalised in the same way as in the published paper on the QSP model of biologics[22].

We described normalised %improved EASI, $m_j(t)$, for the *j*-th drug at *t* by

$$m_j(t) = (m_j^*(t) - m_{\mathrm{P}_j}^*(t)) + m_{\mathrm{P_d}}^*(t), \quad \text{(S2)}$$

where the first term corresponds to the net effects of the drug defined by the difference of the efficacy (%improved EASI) between the *j*-th drug ($m_j^*(t)$) and the corresponding placebo groups ($m_{\mathrm{P}_j}^*(t)$). This term adjusts for different efficacies in the placebo group across the clinical studies due to differences in study participants' background, concomitant drugs and sites of study[21]. The second term corresponds to the placebo effects defined by the efficacy in the placebo group in the dupilumab clinical trial ($m_{\mathrm{P_d}}^*(t)$).

Normalised mean EASI score, $e_j(t)$, of the *j*-th drug at *t* was calculated by

$$e_j(t) = \frac{e_{\mathrm{d}}(0)(100 - m_j(t))}{100}, \quad \text{(S3)}$$

where $e_{\mathrm{d}}(0)$ is the reported baseline (before the trial) mean EASI score in the dupilumab clinical trial[1] and $m_j(t)$ is the normalised %improved EASI defined in (S2).

Normalised EASI-75 was estimated from the normalised %improved EASI using a regression curve obtained from the relationship between %improved EASI and EASI-75 in clinical trials of multiple drugs[12, 15, 16, 17, 18, 19] (FIGURE S3).

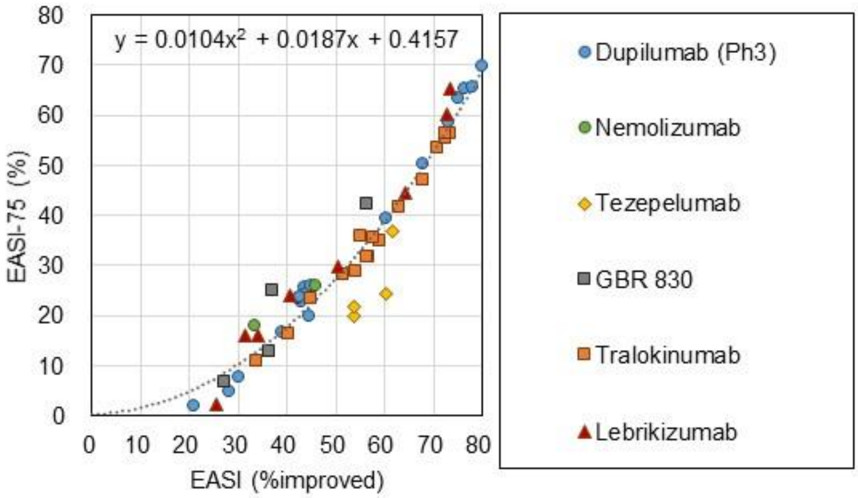


**FIGURE S3** EASI-75 was estimated from %improved EASI using a regression curve. The regression curve was obtained using the reported %improved EASI and EASI-75 in clinical trials of multiple drugs (all available time points of both drug- and placebo-treated groups in dupilumab[12], nemolizumab[15], tezepelumab[16], GBR 830[17], lebrikizumab[18] and tralokinumab[19]).

### 3. Model structure

The QSP model of *S. aureus*-targeted therapies (FIGURE 2) describes the dynamics of EASI score, skin barrier integrity, *S. aureus*, CoNS, IL-4/IL-13 and drug effects. Those dynamics were formulated by Eqs. (S4)-(S21) with six variables (TABLE S2) and 26 parameters (TABLE S3). This section introduces the equations.

*t* is the time after the start of drug treatments. The baseline levels of biological factors for our model (at *t*=0) were obtained from the simulated steady-state level (after 1000 weeks) without any intervention. We referred to the reported levels of biological factors without interventions of the drugs as the reference values for the baseline levels, assuming that the levels of the biological factors were stable before the start of drug treatments.

**TABLE S2** Biological factors as model variables

| Model variables | | Reported baseline levels in AD lesion, Mean (%CV) | | Range |
|---|---|---|---|---|
| $c_4(t)$ | IL-4/IL-13 level at *t* | 39.2 (55) [23 a,c] | Fold change against healthy skin | - |
| $a(t)$ | *S. aureus* level at *t* | 3.4 (43)[20 b] | $Log_{10}$ CFU/cm$^2$ | 0 ~ $a_{max}$ |
| $h(t)$ | CoNS level at *t* | 2.0 (84)[20 b] | $Log_{10}$ CFU/cm$^2$ | 0 ~ $h_{max}$ |
| $a_{agr}(t)$ | Agr expression level at *t* | - [e] | - | 0 (no effect) ~ 1 (maximal effect) |
| $s(t)$ | Skin barrier integrity at *t* | - [e] | - | 0 (complete destruction) ~ 1 (healthy state) |
| $e(t)$ | EASI score at *t* | 29.3 (49)[12 b,c,d] | - | 0 ~ 72 |

a: mild-to-moderate AD patients. Values are average of IL-4 (mean 38.0, %CV 53) and IL-13 (mean 40.5, %CV 56). b: moderate-to-severe AD patients. c: %CV was estimated from IQR. e: no reference data to be compared with simulated values. d: mean baseline value of 29.0 for dupilumab treatment and 29.6 for placebo treatment in dupilumab clinical trial.

**TABLE S3** Model parameters

| Parameters | | Equations | Explored range | | Selected values | |
|---|---|---|---|---|---|---|
| | | | $\mu_i$ | $\sigma_i$ | $\mu_i$ | $\sigma_i$ |
| $k_1$ | Strength of agr expression | S5 | [-2, -1] | [0, 1] | -1.06 | 0.50 |
| $k_2$ | Recovery rate of skin barrier integrity via skin turnover | S6 | [-8, -7] | [0, 1] | -7.71 | 0.33 |
| $k_3$ | Recovery rate of skin barrier integrity via placebo effects | S6 | [-1, 0] | [1, 2] | -0.46 | 1.58 |
| $k_4$ | Proliferation rate of *S. aureus* | S7 | [1, 2] | [0, 1] | 1.37 | 0.20 |
| $k_5$ | Proliferation rate of CoNS | S8 | [-2, -1] | [0, 1] | -1.26 | 0.25 |
| $k_6$ | Secretion rate of IL-4/IL-13 via agr expression | S9 | [-9, -8] | [2, 3] | -8.10 | 2.72 |
| $k_7$ | Secretion rate of IL-4/IL-13 via other pathways | S9 | [-6, -5] | [0, 1] | -5.02 | 0.70 |
| $b_1$ | Inhibitory strength for agr expression via CoNS | S5 | [1, 2] | [0, 1] | 1.37 | 0.04 |
| $b_2$ | Inhibitory strength for recovery of skin barrier via IL-4/IL-13 | S6 | [-3, -2] | [0, 1] | -2.67 | 0.98 |
| $b_3$ | Inhibitory strength for *S. aureus* proliferation via skin barrier | S7 | [-7, -6] | [0, 1] | -6.11 | 0.60 |
| $b_4$ | Inhibitory strength for elimination of Staphylococci via IL-4/IL-13 | S7, S8 | [-3, -2] | [1, 2] | -2.71 | 1.51 |
| $d_1$ | Degradation rate of skin barrier via skin turnover | S6 | [-10, -9] | [1, 2] | -9.86 | 1.41 |
| $d_2$ | Degradation rate of skin barrier via *S. aureus* | S6 | [-9, -8] | [2, 3] | -8.33 | 2.32 |
| $d_3$ | Killing rate of *S. aureus* via bacteriocins secreted from CoNS | S7 | [-5, -4] | [2, 3] | -4.65 | 2.61 |
| $d_4$ | Killing rate of *S. aureus* via AMPs | S7 | [0, 1] | [0, 1] | 0.55 | 0.39 |
| $d_5$ | Elimination rate of *S. aureus* via turnover | S7 | [0, 1] | [0, 1] | 0.23 | 0.19 |
| $d_6$ | Killing rate of CoNS via bacteriocins secreted from *S. aureus* | S8 | [-9, -8] | [0, 1] | -8.14 | 0.59 |
| $d_7$ | Killing rate of CoNS via AMPs | S8 | [-4, -3] | [1, 2] | -3.44 | 1.88 |
| $d_8$ | Elimination rate of CoNS via turnover | S8 | [-2, -1] | [0, 1] | -1.73 | 0.40 |
| $d_9$ | Elimination rate of IL-4/IL-13 | S9 | [-9, -8] | [1, 2] | -8.62 | 1.19 |
| $d_{A9a\_s}$ | Killing rate of *S. aureus* via *Sh*A9 in bacteriocin-sensitive *S. aureus* | S13 | [1, 2] | [0, 1] | 1.09 | 0.90 |
| $d_{A9a\_r}$ | Killing rate of *S. aureus* via *Sh*A9 in bacteriocin-resistant *S. aureus* | S13 | [-1, 0] | [0, 1] | -0.83 | 0.85 |
| $d_{A9h}$ | Killing rate of CoNS via *Sh*A9 | S11 | [0, 1] | [0, 1] | 0.55 | 0.90 |
| $b_{A9s}$ | Inhibitory strength for agr expression via *Sh*A9 | S12 | [-2, -1] | [0, 1] | -1.11 | 0.27 |
| $d_{fs}$ | Killing rate of S. aureus via flucloxacillin | S14 | [0, 1] | [0, 1] | 0.13 | 0.27 |
| $d_{fh}$ | Killing rate of CoNS via flucloxacillin | S15 | [0, 1] | [0, 1] | 0.35 | 0.12 |

#### 3.1. Biological factors

(a) Agr expression level

Agr expression level, $a_{\mathrm{agr}}(t)$, of *S. aureus* is described (FIGURE S4) by

$$a_{\mathrm{agr}}(t) = \tanh \frac{k_1 a(t)}{1+b_1 h(t)}, \tag{S4}$$

where $a(t)$ and $h(t)$ are *S. aureus* level and CoNS level ($\log_{10}$ CFU/$cm^2$), respectively and $b_1$ describes the inhibitory strength for the agr expression by AIPs from CoNS[24].

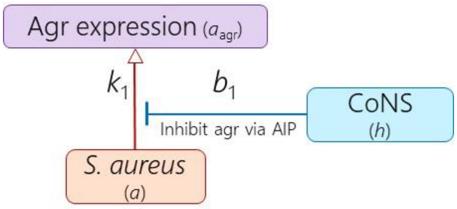


**FIGURE S4** Agr expression is regulated by *S. aureus* and CoNS

(b) Skin barrier integrity

The dynamics of the skin barrier integrity, $s(t)$, is described (FIGURE S5) by

$$\frac{ds(t)}{dt} = \frac{(1-s(t))(k_2 + k_3)}{1+b_2 c_4(t)} - s(t)\{d_1 + d_2 a_{\mathrm{agr}}(t)\}, \tag{S5}$$

where $c_4(t)$ is IL-4/IL-13 level, $s(t)$ is skin barrier integrity, $k_2$ and $k_3$ describe the recovery rate of skin barrier integrity via skin turnover and that via placebo effects, respectively, $b_2$ describes the inhibitory strength for recovery of skin barrier via IL-4/IL-13 and $d_1$ and $d_2$ describe the degradation rate of skin barrier via skin turnover and that via *S. aureus*, respectively.

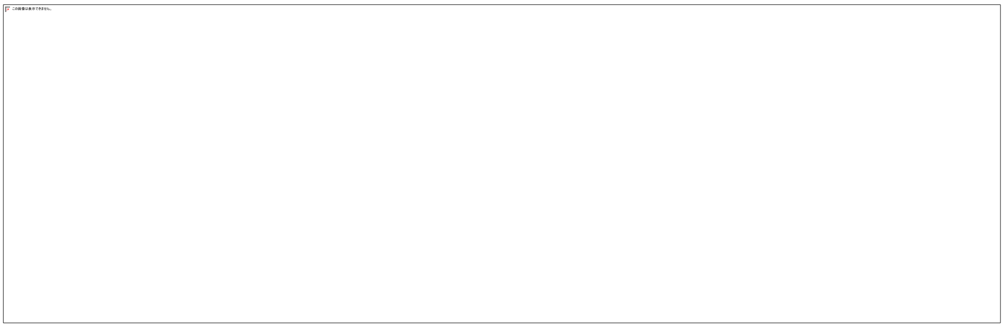

**FIGURE S5** Skin barrier integrity is regulated by skin turnover, placebo effects, IL-4/13 and agr expression. Squared and oval symbols represent model variables and implicit factors in our model, respectively.

The first term represents a recovery of skin barrier integrity by intrinsic skin turnover (with the recovery rate, $k_2$) and placebo effects ($k_3$). We assumed the maximal value of $s(t) = 1$ as a healthy state of skin barrier integrity (TABLE S2), and thus modified the recovery rate by $1 - s(t)$. The placebo effect was applied to the simulations for both placebo- and drug-treated groups, as placebo-treated patients improved the EASI score[11,12,20], presumably because of the controlled care with concomitant drugs such as emollients during the clinical trials. The recovery of skin barrier integrity was assumed to be compromised by IL-4[25] and IL-13[26] (with the strength $b_2$) as they are shown to decrease filaggrin production and inhibit epidermal differentiation.

The second term corresponds to the degradation of the skin barrier by skin turnover (with the degradation rate, $d_1$) and by *S. aureus,* which damages keratinocytes through phenol-soluble modulin-α (PSMα) and δ-toxin ($d_2$)[27]. The latter ($d_2$) is agr-dependent because the agr regulates secretion of PSMα and δ-toxin from *S. aureus*[28].

(c) *S. aureus* and CoNS in the skin

The dynamics of *S. aureus* and CoNS in the skin, $a(t)$ and $h(t)$, are described (FIGURE S6) by

$$\frac{da(t)}{dt} = \frac{k_4}{1+b_3 s(t)}\left(1 - \frac{a(t)}{a_{\max}}\right) - \left\{d_3 h(t) + \frac{d_4}{1+b_4 c_4(t)} + d_5\right\} \text{ and} \quad \text{(S6)}$$

$$\frac{dh(t)}{dt} = k_5\left(1 - \frac{h(t)}{h_{\max}}\right) - \left\{d_6 a(t) + \frac{d_7}{1+b_4 c_4(t)} + d_8\right\}, \quad \text{(S7)}$$

where $k_4$ and $k_5$ are the proliferation rates of *S. aureus* and CoNS, respectively, $b_3$ is the inhibitory coefficient for *S. aureus* proliferation via skin barrier, $b_4$ is the inhibitory strength for elimination of Staphylococci via IL-4/IL-13, $d_3$ and $d_4$ are the killing rate of *S. aureus* via bacteriocins secreted from CoNS and that via AMPs, respectively, $d_5$ is the elimination rate of *S. aureus* via turnover, $d_6$ and $d_7$ are the killing rate of CoNS via bacteriocins secreted from *S. aureus* and that via AMPs, respectively, $d_8$ is the elimination rate of CoNS via turnover and $a_{\max}$ and $h_{\max}$ are the maximal levels of *S. aureus* and CoNS, respectively. Eq.(S6 and S7) represent logistic growth of $a(t)$ and $h(t)$ in $\log_{10}$ scale. We set $[a_{\max}, h_{\max}] = [7, 7]$ to cover the reported range of *S. aureus* levels (reported maximal $\log_{10}$ level of *S. aureus* was 6) in the dupilumab clinical trial[11].

The Eqs (S6 and7) are relative growth rates based on $\log_{10}$ scale ($\log_{10}$ CFU/cm$^2$). Their absolute gowth rates can be described as

$$a^*(t) = 10^{a(t)} \quad \text{(S8)}$$

$$h^*(t) = 10^{h(t)} \quad \text{(S9)}$$

$$\frac{da^*(t)}{dt} = \frac{da(t)}{dt} a^*(t) \ln 10$$
$$= \frac{k_4}{1+b_3 s(t)}\left(1 - \frac{\log_{10} a^*(t)}{a_{\max}}\right) a^*(t) \ln 10 - \left\{d_3 \log_{10} h^*(t) + \frac{d_4}{1+b_4 c_4(t)} + d_5\right\} a^*(t) \ln 10 \quad \text{(S10)}$$

$$\frac{dh^*(t)}{dt} = \frac{dh(t)}{dt} h^*(t) \ln 10$$
$$= k_5\left(1 - \frac{\log_{10} h^*(t)}{h_{\max}}\right) h^*(t) \ln 10 - \left\{d_6 \log_{10} a^*(t) + \frac{d_7}{1+b_4 c_4(t)} + d_8\right\} h^*(t) \ln 10 \quad \text{(S11)}$$

where $a^*(t)$ and $h^*(t)$ are absolute levels (CFU/cm$^2$) of *S. aureus* and CoNS in the skin, respectively. The first terms of Eqs.(S10 and 11) mean that we assumed that their logistic growth is based on log10 scale of *S. aureus* and CoNS levels.

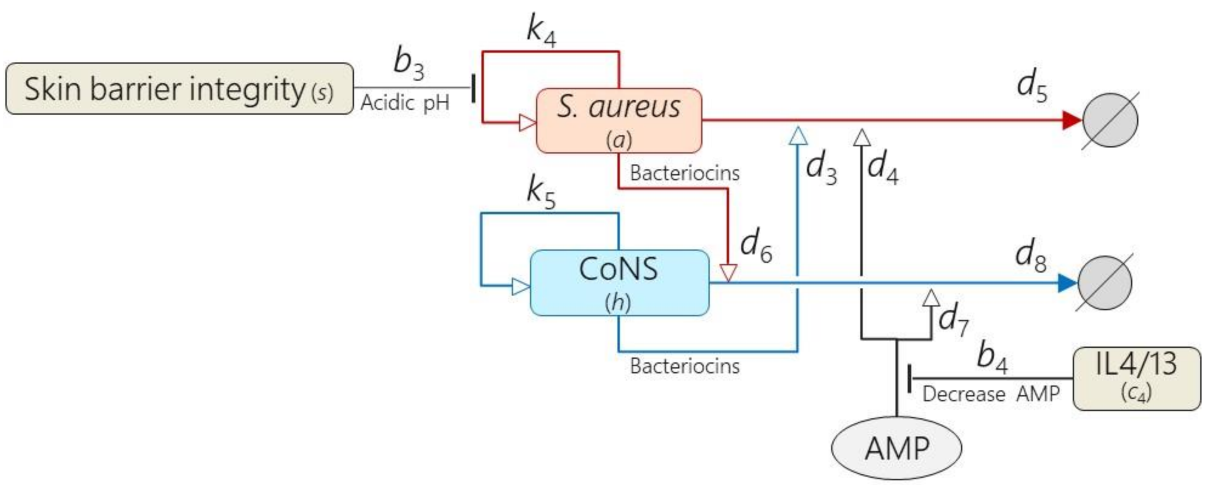


**FIGURE S6** *S. aureus* and CoNS levels regulate each other. Squared and oval symbols represent model variables and implicit factors in our model, respectively.

*S. aureus* and CoNS proliferate (with the rates $k_4$ and $k_5$), where healthy skin barrier integrity inhibits proliferation of *S. aureus* by making skin pH acidic (with strength $b_3$) whereas skin pH does not affect those of CoNS[29, 30].

*S. aureus* and CoNS are killed by bacteriocins (released from Staphylococci) and AMP (released from keratinocytes) directly[31]. Bacteriocins exert antimicrobial activity against bacteria closely related to the producer strain but not against the producer strain itself[32]; *S aureus* is killed by bacteriocins from CoNS (with strength $d_3$)[33] and AMP ($d_4$), and CoNS is killed by bacteriocins from *S. aureus* ($d_6$)[34] and AMP ($d_7$). AMP release from keratinocytes is inhibited by IL-4 and IL-13[35] ($b_4$). *S. aureus* and CoNS in the skin decrease due to their natural death ($d_5$ and $d_8$).

We did not consider influence of *S. aureus* on AMP because the experimental evidence is controversial: *S. aureus* increases AMP release from keratinocytes via pathways that are independent of the cytokines[36]; *S. aureus* degrades AMP by aureolysin, which is a proteinase produced by *S. aureus*[37].

(d) IL-4 and IL-13

The dynamics of the IL-4 and IL-13, $c_4(t)$, is described (FIGURE S7) by

$$\frac{dc_4(t)}{dt} = k_6 a_{\text{agr}}(t) + k_7 - d_9 c_4(t), \tag{S12}$$

where $k_6$ and $k_7$ are the secretion rate of IL-4/IL-13 via agr expression and that via other pathways, respectively and $d_9$ is the elimination rate of IL-4/IL-13.

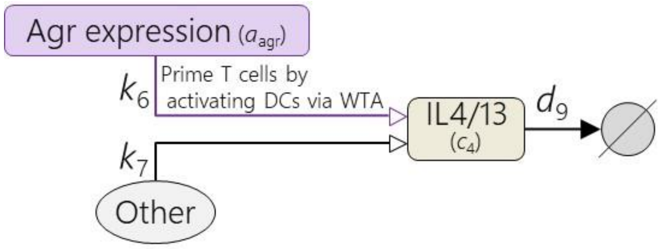


**FIGURE S7** IL-4/IL-13 level is regulated by agr expression and other factors. Squared and oval symbols represent model variables and implicit factors in our model, respectively.

IL-4 and IL-13 are secreted from Th2 cells that are primed by dendritic cells (DCs) specifically activated by *S. aureus*-derived wall teichoic acid (WTA)[38] controlled by agr[39] (with the rate $k_6$). There are other pathways releasing IL-4/IL-13, which were implicitly described as "other" effects ($k_7$).

(e) EASI score

The EASI score (ranging from 0 to 72) is calculated using the severity and the area scores of equally-weighted four AD signs (erythema, induration, excoriations and lichenification) on four body regions (head/neck, trunk, upper limbs and lower limbs)[40]. In our model, the EASI score, $e(t)$, is described (FIGURE S8) by

$$e(t) = 72\,\frac{2a_{\mathrm{agr}}(t)+2(1-s(t))}{4}, \tag{S13}$$

where 72 is the maximal EASI score. Scores derived from two AD signs (erythema and induration) and those from the remaining two signs (excoriations and lichenification) were surrogated by $a_{\mathrm{agr}}(t)$ and $1-s(t)$, respectively, as described below. We set $e(0) = 29.3$, the baseline EASI score of the AD patients in the dupilumab clinical trial, which was used as a reference value to normalise the EASI scores in all the clinical trials.

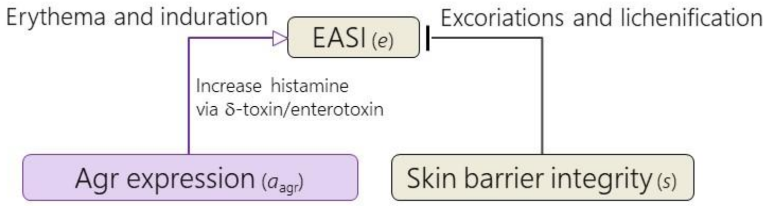


**FIGURE S8** EASI score was calculated from agr expression and skin barrier integrity

We assumed the scores derived from erythema and induration are governed by $a_{\mathrm{agr}}(t)$ because these two signs can be induced by *S. aureus*[41]. We used $\log_{10}$ level of *S. aureus* to model $a_{\mathrm{agr}}(t)$ in Eq. (S4) as the correlation between EASI score and $\log_{10}$ level of *S. aureus* has been reported[11]. Erythema is caused by inflammatory vasodilation by histamines[42]. Histamine is released mainly from mast cells and basophils that are activated by detecting antigens, such as δ-toxin[43] and Staphylococcus enterotoxins[44], released by *S. aureus* but not by CoNS[45]. We associated the released histamine concentration with the antigen load in this model because the amount of histamine release depends more on the amount of antigens than that of antigen-specific IgE[46], although both antigens and antigen-specific IgE play a role in this process[47]. A negligible contribution of IgE (compared to that of antigens) on the AD pathogenesis is also suggested by a lack of clinical efficacy demonstrated for omalizumab (IgE neutralizing anti-IgE antibody). Our model assumed that the histamine release by *S. aureus*-induced δ-toxin and enterotoxins depends on the agr expression level of *S. aureus* because AIPs from other strains regulate secretion of δ-toxin and enterotoxins from *S. aureus*[28,48].

Scores for the other two AD signs, excoriations and lichenification, are surrogated by $1-s(t)$, which describes the degree of damage of the skin barrier integrity, because excoriations and lichenification are caused by scratching[49], which damages skin barrier integrity.

### 3.2. Drug effects

#### 3.2.1. Flucloxacillin

Flucloxacillin, an antibiotic, kills the Staphylococci (*S. aureus* and CoNS). We described the effects of flucloxacillin (FIGURE S9) on decreasing the Staphylococci by adding the killing rates of Staphylococci ($d_{\mathrm{fa}}$ and $d_{\mathrm{fh}}$) in Eq.(S6 and S7):

$$\frac{da(t)}{dt} = \frac{k_4}{1+b_3 s(t)}\left(1 - \frac{a(t)}{a_{\max}}\right) - \left\{d_4 h(t) + \frac{d_5}{1+b_4 c_4(t)} + d_6 + d_{\mathrm{fa}}\right\}, \quad \text{(S14)}$$

$$\frac{dh(t)}{dt} = k_5\left(1 - \frac{h(t)}{h_{\max}}\right) - \left\{d_6 a(t) + \frac{d_7}{1+b_4 c_4(t)} + d_8 + d_{\mathrm{fh}}\right\}. \quad \text{(S15)}$$

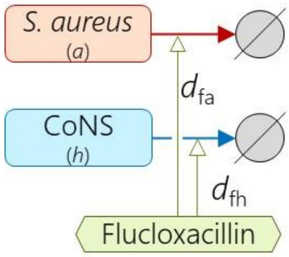


**FIGURE S9** Effects of flucloxacillin. Squared and hexagon symbols represent model variables and a drug in our model, respectively.

#### 3.2.2. *S. hominis* A9 (*Sh*A9)

*Sh*A9 is a specific strain of *S. hominis*, which produces bacteriocins against *S. aureus*[20] and inhibits agr expression of *S. aureus*[24]. Although *Sh*A9 was screened based on selectivity of the bacteriocins against *S. aureus*, it still has antimicrobial activity against CoNS[33]. We describe those effects (FIGURE S10) by adding the killing rates of *S. aureus* and CoNS ($d_{\mathrm{A9a}}$ and $d_{\mathrm{A9h}}$) in Eq.(S6 and S7) and the inhibitory strength for agr expression ($b_{\mathrm{A9a}}$) in Eq.(S4):

$$\frac{da(t)}{dt} = \frac{k_4}{1+b_3 s(t)}\left(1 - \frac{a(t)}{a_{\max}}\right) - \left\{d_4 h(t) + \frac{d_5}{1+b_4 c_4(t)} + d_6 + d_{\mathrm{A9a}}\right\}, \quad \text{(S16)}$$

$$\frac{dh(t)}{dt} = k_5\left(1 - \frac{h(t)}{h_{\max}}\right) - \left\{d_6 a(t) + \frac{d_7}{1+b_4 c_4(t)} + d_8 + d_{\mathrm{A9h}}\right\}, \quad \text{(S17)}$$

$$a_{\mathrm{agr}}(t) = \tanh \frac{k_1 a(t)}{(1+b_1 h(t))(1+b_{\mathrm{A9a}})}. \quad \text{(S18)}$$

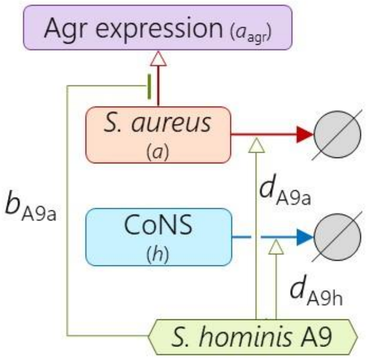


**FIGURE S10** Effects of *S. hominis* A9. Squared and hexagon symbols represent model variables and a drug in our model, respectively.

The clinical trial of *Sh*A9 stratified the patients according to the sensitivity of *S. aureus* isolated from each patient to the bacteriocins of *Sh*A9. The colonised *S. aureus* was categorised as "sensitive" when minimal inhibitory concentration (MIC) of *Sh*A9 conditioned medium against *S. aureus* is less than 100% (% of original conditioned medium) and as "resistant" when the MIC is more than 200%[20]. Hereafter, *Sh*A9 applied to patients colonised with *S. aureus* that is sensitive to *Sh*A9 bacteriocins is referred as *Sh*A9-sensitive and those with *S. aureus* that is resistant to *Sh*A9 bacteriocins is referred as *Sh*A9-resistant. We modelled the different sensitivity of *S. aureus* to the bacteriocins of *Sh*A9 as

$$d_{\mathrm{A9a}} = \begin{cases} d_{\mathrm{A9a\_s}}, & \text{if } Sh\mathrm{A9} - \text{sensitive} \\ d_{\mathrm{A9a\_r}}, & \text{if } Sh\mathrm{A9} - \text{resistant}. \end{cases} \quad \text{(S13)}$$

Effects of *Sh*A9 were applied in both dosing and follow-up periods in the simulation because the measured amount of *Sh*A9 remained higher than baseline levels during the follow-up periods in the clinical trial[20].

#### 3.2.3. Dupilumab

We described the effects of dupilumab (FIGURE S11) that inhibit the signalling of IL-4 and IL-13 by scaling the concentrations of IL-4 and IL-13. Effective concentrations of the IL-4 and IL-13 in the skin at *t*, $c_4(t)$, was modelled by

$$c_4(t) = (1 - r_{\mathrm{inhibit}})c_4^*(t), \quad \text{(S19)}$$

$$r_{\mathrm{inhibit}} = \frac{d_{\mathrm{skin}}}{\mathrm{IC}_{50} + d_{\mathrm{skin}}}, \quad \text{(S20)}$$

$$d_{\mathrm{skin}} = r_{\mathrm{skin/serum}} d_{\mathrm{serum}}, \quad \text{(S21)}$$

where $c_4^*(t)$ is the concentration of IL-4 and IL-13 in the skin at *t*, $r_{\mathrm{inhibit}}$ is the rate of the IL-4 and IL-13 inhibition in the dupilumab treatment, $d_{\mathrm{skin}}$ is the concentration of dupilumab in the skin, $\mathrm{IC}_{50}$ is the half-maximal inhibitory concentration of dupilumab against IL-4 and IL-13, $r_{\mathrm{skin/serum}}$ is the ratio of dupilumab concentration in the skin to that in serum and $d_{\mathrm{serum}}$ is the mean concentration of the dupilumab in serum. We adopted $r_{\mathrm{skin/serum}} = 0.157$ for dupilumab based on the estimated ratio of antibody concentration in the skin to that in the plasma[50]. Values of $\mathrm{IC}_{50}$ (IL-4: <0.01 and IL-13: 0.01 mcg/mL) and $d_{\mathrm{serum}}$ (183 mcg/mL) were obtained from reported results of in vitro assay and the reported pharmacokinetic data of the adopted dose regimen (TABLE 1) in clinical trials[51, 52]. With these values, $r_{\mathrm{inhibit}}$ was calculated as 0.99.

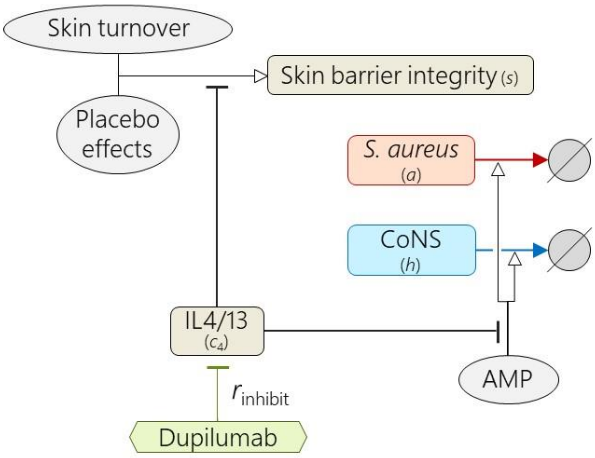


**FIGURE S11** Effects of dupilumab. Squared, oval and hexagon symbols represent model variables, implicit factors and a drug in our model, respectively.

### 4. Optimising model parameters to reproduce clinical data

We optimised 52 parameters ($\mu_i$ and $\sigma_i$) that define the distributions of the 26 model parameters (TABLE S3) so that the model reproduces the following clinical data consisting of 108 reference values;

- mean values and %CV of 4 biological factors (IL-4/IL-13, *S. aureus*, CoNS and the EASI score) without interventions of the drugs (TABLE S2; 2 indices x 4 factors = 8 reference values),
- the EASI score and EASI-75 in the clinical trials (FIGURE 1; 2 indices x 5 interventions x 4-7 time points/intervention = 56 reference values) and
- mean values and %CV of *S. aureus* levels in the clinical trials (FIGURE 1; 2 indices x 5 interventions x 4-5 time points/intervention = 44 reference values).

We searched the parameters that minimize the cost function, *J*, defined by

$$J = w_1 J_1 + w_2 J_2 + w_3 J_3 + w_4 J_4 + w_5 J_5 + w_6 J_6, \quad \text{(S22)}$$

where

$$J_1 = \sqrt{\frac{1}{4}\sum_{l=1}^{4}\left(b_{\mathrm{mean},l} - \hat{b}_{\mathrm{mean},l}\right)^2}, \quad \text{(S23)}$$

$$J_2 = \sqrt{\frac{1}{4}\sum_{l=1}^{4}\left(b_{\mathrm{CV},l} - \hat{b}_{\mathrm{CV},l}\right)^2}, \quad \text{(S24)}$$

$$J_3 = \sqrt{\frac{1}{5}\sum_{j=1}^{5}\left\{\frac{1}{m_{\mathrm{last},j}}\sum_{m=1}^{m_{\mathrm{last},j}}\left(e_j(t_m) - \hat{e}_j(t_m)\right)^2\right\}}, \quad \text{(S25)}$$

$$J_4 = \sqrt{\frac{1}{5}\sum_{j=1}^{5}\left\{\frac{1}{m_{\mathrm{last},j}}\sum_{m=1}^{m_{\mathrm{last},j}}\left(e_{75,j}(t_m) - \hat{e}_{75,j}(t_m)\right)^2\right\}}. \quad \text{(S26)}$$

$$J_5 = \sqrt{\frac{1}{5}\sum_{j=1}^{5}\left\{\frac{1}{m_{\mathrm{last},j}}\sum_{m=1}^{m_{\mathrm{last},j}}\left(a_j(t_m) - \hat{a}_j(t_m)\right)^2\right\}}, \quad \text{(S27)}$$

$$J_6 = \sqrt{\frac{1}{5}\sum_{j=1}^{5}\left\{\frac{1}{m_{\mathrm{last},j}}\sum_{m=1}^{m_{\mathrm{last},j}}\left(a_{\mathrm{CV},j}(t_m) - \hat{a}_{\mathrm{CV},j}(t_m)\right)^2\right\}}. \quad \text{(S28)}$$

The terms, $J_1$ and $J_2$, are root mean squared errors (RMSE) of mean values and %CV of baseline levels of biological factors, respectively, $J_3$ and $J_4$ are RMSE of the EASI score and EASI-75, respectively, $J_5$ and $J_6$ are RMSE of mean values and %CV of *S. aureus* levels, respectively. $w_1$ to $w_6$ are the weighting coefficients. $b_{\mathrm{mean},l}$ and $b_{\mathrm{CV},l}$ are the reference values for the mean value and the %CV of baseline levels of the $l$-th biological factor ($l$=1,2,3,4). $\hat{b}_{\mathrm{mean},l}$ and $\hat{b}_{\mathrm{CV},l}$ are the corresponding simulated values at the steady state (after 1000 weeks, among 1000 virtual patients). $e_j(t_m)$, $e_{75,j}(t_m)$, $a_j(t_m)$ and $a_{\mathrm{CV},j}(t_m)$ are the reference values of the EASI score, EASI-75, mean *S. aureus* levels and %CV of *S. aureus* levels using the $j$-th intervention ($j$=1,2,3,4,5) at time $t_m$ ($m$=1,…, $m_{\mathrm{last},j}$). $\hat{e}_j(t_m)$, $\hat{e}_{75,j}(t_m)$ , $\hat{a}_j(t_m)$ and $\hat{a}_{\mathrm{CV},j}(t_m)$ are the corresponding simulated values. We used $[w_1, w_2, w_3, w_4, w_5, w_6] = [50, 10, 50, 50, 1000, 1]$ with larger weights on some terms (e.g., $J_5$) that tended to have smaller fitting errors.

The parameters were optimised using differential evolution[53], which is an effective method for global optimisation of a large number of parameters. The conditions for differential evolution were set as follows based on manual trial-and-error.

| | |
|---|---|
| Mutation constant (F) | : 0.5 |
| Crossover constant (CR) | : 0.7 |
| Strategy | : DE/best/1/bin |
| Number of population vectors (NP) | : 52 |
| Number of function evaluations (nfe) | : 15652 |
| Number of evaluated generations | : 300 |
| Ranges of parameters searched | : TABLE S3 |

The *J* reached a plateau value, 569, after the iterative evaluations (FIGURE S12). The model fitness was confirmed visually by comparing the reference and simulated data (FIGURE 3).

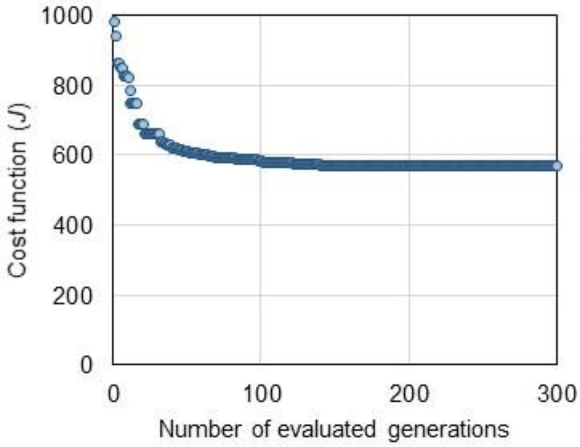


**FIGURE S12** The cost function (*J*) reached a plateau value, 569, in the optimisation process using differential evolution.

### 5. Sensitivity analysis

We conducted a global sensitivity analysis of the model parameters with respect to %improved EASI. We produced 1000 virtual patients by varying the 26 parameters that represent their pathophysiological backgrounds using Latin hypercube sampling (LHS) and computed partial rank correlation coefficient (PRCC)[54] between each parameter and %improved EASI of each drug. LHS is a sampling method to explore the entire space of multidimensional parameters efficiently, and PRCC represents a rank correlation coefficient that is controlled for confounding effects that could lead to detecting pseudo-correlations. The evaluated ranges of $\ln k_i$ were $[\mu_i - \sigma_i, \mu_i + \sigma_i]$. The *p*-values for the PRCC were adjusted for multiple testing with the Bonferroni procedure, where a significance level of adjusted $p < 0.05$ with an absolute value >0.1 was used.

#### 5.1. Influence of model parameters on efficacy of placebo

Eight model parameters had a significant PRCC with the %improved EASI by placebo (FIGURE S13).

Two out of the eight parameters were skin barrier-related ($k_3$ and $b_2$). A higher $k_3$ results in stronger recovery of skin barrier via placebo effects, thereby achieving a higher %improve EASI. A higher $b_2$ inhibits recovery of skin barrier more strongly, weakening the recovery of the skin barrier by placebo effects, thereby showing lower %improve EASI.

The remaining six parameters were agr-related ($k_1$, $k_4$, $k_5$, $b_1$, $d_5$ and $d_8$). Higher $b_1$, $k_5$ and $d_5$ and lower $k_1$, $k_4$ and $d_8$ result in a lower baseline level of agr expression via decreasing agr expression levels ($k_1$ and $b_1$) and *S. aureus* levels ($k_4$ and $d_5$) or increasing CoNS levels ($k_5$ and $d_8$). A lower level of agr expression means that the % improved EASI score is more sensitive to the changes in the skin barrier integrity that is achieved by placebo effects because we modelled an EASI score as a weighted mean of agr expression and skin barrier integrity (Eq. S4).

These influences were observed in not only placebo groups but also drug-treated groups as the placebo effects were considered in both placebo- and drug-treated groups in the simulation.

#### 5.2. Influence of model parameters on efficacy of dupilumab

Six model parameters had a significant PRCC with the %improved EASI by dupilumab (FIGURE S13).

All the six parameters were agr-related ($k_1$, $k_4$, $k_5$, $d_4$, $d_5$ and $d_8$). Higher $k_5$, $d_4$ and $d_5$ and the lower $k_1$, $k_4$ and $d_8$ result in a lower baseline level of agr expression due to a decrease in agr expression ($k_1$) and in *S. aureus* levels ($k_4$, $d_4$ and $d_5$) or an increase in CoNS levels ($k_5$ and $d_8$). A lower level of agr expression means that the % improved EASI score is more sensitive to the changes in the skin barrier integrity that is achieved by placebo effects and dupilumab (inhibiting skin barrier damage from IL-4/IL-13) because we modelled an EASI score as a weighted mean of agr expression and skin barrier integrity (Eq. S4).

Two skin barrier-related parameters ($k_3$ and $b_2$) had a significant PRCC with the %improved EASI by placebo, but not by dupilumab, which includes placebo effects in our simulation. It may be because the recovery of the skin barrier by dupilumab overweighted that by placebo effects, and thereby the placebo effects became negligible in dupilumab treatment.

#### 5.3. Influence of model parameters on efficacy of *Sh*A9-sensitive

Seven model parameters had a significant PRCC with the %improved EASI by *Sh*A9-sensitive (FIGURE S13).

Two out of the seven parameters were skin barrier-related ($k_3$ and $b_2$) and correspond to placebo effects because they had a significant PRCC with the %improved EASI by placebo (Section SI 5.1). Other two parameters were bactericidal strengths of *Sh*A9 ($d_{A9a\_s}$ and $d_{A9h}$). A higher $d_{A9a\_s}$ and a lower $d_{A9h}$ result in stronger killing of *S. aureus* and weaker killing of CoNS, thereby achieving a higher %improve EASI.

The remaining three parameters were agr-related ($k_4$, $k_5$, $d_8$). As described in SI 5.1, a lower $k_4$ showed a higher %improve EASI in placebo treatment. A higher $d_8$ and a lower $k_5$ result in a lower baseline level of CoNS. The lower level of CoNS lessens the impact of killing CoNS by *Sh*A9 on the increase of agr expression. The smaller increase in agr expression results in the weaker detrimental effects of *Sh*A9 on EASI scores, thereby showing a higher %improve EASI.

The influences of $d_8$ and $k_5$ (i.e., a baseline level of CoNS) on %improve EASI were in opposite directions, depending on whether the drugs kill CoNS (e.g., *Sh*A9 and flucloxacillin) or not (e.g., placebo and dupilumab). A higher baseline level of CoNS makes a lower baseline level of agr expression. The lower level of agr expression means that the %improved EASI score is more sensitive to the changes in the skin barrier integrity that is achieved by placebo and dupilumab because we modelled an EASI score as a weighted mean of agr expression and skin barrier integrity (Eq. S4). On the other hand, a lower level of CoNS lessens the impact of killing CoNS by *Sh*A9 on the increase of agr expression. The smaller increase in agr expression results in the weaker detrimental effects of *Sh*A9 and flucloxacillin on EASI scores, thereby showing a higher %improve EASI.

$b_{A9s}$ (inhibitory strength for agr expression of *S. aureus* via *Sh*A9) had no significant influence on %improve EASI (FIGURE S13) because the inhibitory strength of *Sh*A9 for agr expression of *S. aureus* is so weak in this model (i.e., a small $\mu_i$ of $b_{A9s}$) that the sensitivity analysis evaluated a narrow range of inhibition levels of agr expression (The evaluated range of $b_{A9s}$ was 0.25-0.43, 20%-30% around the nominal value). On the other hand, the inhibitory level of agr expression via hypothetical *S. aureus*-targeted therapy had a significant influence on EASI-75 (FIGURE 5) because it evaluated a whole range of inhibition levels of agr expression (0%-100%).

### 5.4. Influence of model parameters on efficacy of *Sh*A9-resistant

Six model parameters had a significant PRCC with the %improved EASI by *Sh*A9-resistant (FIGURE S13).

Two out of the six parameters were skin barrier-related ($k_3$ and $b_2$) and correspond to placebo effects because they had a significant PRCC with the %improved EASI by placebo (Section SI 5.1). A parameter, $d_{A9h}$, is the bactericidal strength of *Sh*A9 on CoNS. A lower $d_{A9h}$ results in weaker killing of CoNS, thereby achieving a higher %improve EASI.

The remaining three parameters were agr-related ($k_4$, $d_5$ and $d_8$). As described in SI 5.1, a higher $d_5$ and a lower $k_4$ result in stronger skin barrier recovery by placebo effects, and thereby showed a higher %improve EASI. A higher $d_8$ result in a lower baseline level of CoNS. The lower level of CoNS lessens the impact of CoNS killing by *Sh*A9 on the increase of agr expression. The smaller increase in agr expression results in weaker detrimental effects of *Sh*A9 on EASI scores, thereby showing a higher %improve EASI.

*Sh*A9-resistant and *Sh*A9-sensitive showed similar results except for $k_5$, $d_5$, $d_{A9a\_s}$/$d_{A9a\_r}$; the discrepancy stems from the difference in bactericidal strengths on *S. aureus*.

### 5.5. Influence of model parameters on efficacy of flucloxacillin

Eight model parameters had a significant PRCC with the %improved EASI by flucloxacillin (FIGURE S13).

Two out of the eight parameters were skin barrier-related ($k_3$ and $b_2$) and correspond to placebo effects (Section SI 5.1). Other two parameters were bactericidal strengths of flucloxacillin ($d_{fa}$ and $d_{fh}$). A higher $d_{fa}$ and a lower $d_{fh}$ result in stronger killing of *S. aureus* and weaker killing of CoNS, thereby achieving a higher %improve EASI.

The remaining four parameters were agr-related ($k_4$, $k_5$, $d_5$ and $d_8$). As described in SI 5.1, a higher $d_5$ and a lower $k_4$ result in stronger recovery of skin barrier via placebo effects, thereby showing a higher %improve EASI. A higher $d_8$ and a lower $k_5$ have a lower baseline level of CoNS. The lower level of CoNS lessens the impact of killing CoNS by flucloxacillin on the increase of agr expression. The smaller increase in agr expression results in weaker detrimental effects of *Sh*A9 on EASI scores, thereby showing a higher %improve EASI.

| Parameters | | %improved EASI: Placebo | Dupilumab | *Sh* A9-sensitive | *Sh* A9-resistant | Flucloxacillin |
|---|---|---|---|---|---|---|
| Strength of agr expression of *S. aureus* | k1 | -0.3 | -0.2 | ~~0.0~~ | ~~-0.1~~ | ~~-0.1~~ |
| Recovery rate of skin barrier integrity via skin turnover | k2 | ~~0.0~~ | ~~0.0~~ | ~~0.0~~ | ~~0.0~~ | ~~0.0~~ |
| Recovery rate of skin barrier integrity via placebo effects | k3 | 0.5 | ~~0.1~~ | 0.4 | 0.2 | 0.3 |
| Proliferation rate of *S. aureus* | k4 | -0.5 | -0.7 | -0.3 | -0.4 | -0.6 |
| Proliferation rate of CoNS | k5 | 0.5 | 0.4 | -0.2 | ~~-0.1~~ | -0.3 |
| Secretion rate of IL-4/IL-13 via agr expression | k6 | ~~0.0~~ | ~~0.0~~ | ~~0.0~~ | ~~0.0~~ | ~~0.0~~ |
| Secretion rate of IL-4/IL-13 via other pathways | k7 | ~~-0.1~~ | ~~0.0~~ | ~~0.0~~ | ~~0.0~~ | ~~0.0~~ |
| Inhibitory strength for agr expression via CoNS | b1 | 0.2 | ~~0.1~~ | ~~-0.1~~ | ~~-0.1~~ | ~~-0.1~~ |
| Inhibitory strength for recovery of skin barrier via IL-4/IL-13 | b2 | -0.4 | ~~0.0~~ | -0.3 | -0.2 | -0.2 |
| Inhibitory strength for *S. aureus* proliferation via skin barrier | b3 | ~~0.0~~ | ~~0.0~~ | ~~0.0~~ | ~~0.0~~ | ~~-0.1~~ |
| Inhibitory strength for elimination of Staphylococci via IL-4/IL-13 | b4 | ~~-0.1~~ | ~~0.0~~ | ~~0.0~~ | ~~0.0~~ | ~~-0.1~~ |
| Degradation rate of skin barrier via skin turnover | d1 | ~~0.0~~ | ~~0.0~~ | ~~0.0~~ | ~~0.1~~ | ~~0.0~~ |
| Degradation rate of skin barrier via *S. aureus* | d2 | ~~0.0~~ | ~~0.0~~ | ~~0.0~~ | ~~0.0~~ | ~~0.1~~ |
| Killing rate of *S. aureus* via bacteriocins secreted from CoNS | d3 | ~~0.0~~ | ~~0.0~~ | ~~0.0~~ | ~~0.0~~ | ~~0.0~~ |
| Killing rate of *S. aureus* via AMPs | d4 | ~~0.1~~ | 0.5 | ~~0.0~~ | ~~0.1~~ | ~~0.1~~ |
| Elimination rate of *S. aureus* via turnover | d5 | 0.4 | 0.3 | ~~0.0~~ | 0.2 | 0.2 |
| Killing rate of CoNS via bacteriocins secreted from *S. aureus* | d6 | ~~0.0~~ | ~~0.0~~ | ~~-0.1~~ | ~~0.0~~ | ~~0.0~~ |
| Killing rate of CoNS via AMPs | d7 | ~~-0.1~~ | ~~-0.1~~ | ~~0.1~~ | ~~0.1~~ | ~~0.0~~ |
| Elimination rate of CoNS via turnover | d8 | -0.5 | -0.4 | 0.2 | 0.2 | 0.3 |
| Elimination rate of IL-4/IL-13 | d9 | ~~0.1~~ | ~~0.0~~ | ~~0.1~~ | ~~0.1~~ | ~~0.1~~ |
| Killing rate of *S. aureus* via *Sh* A9 in bacteriocin-sensitive *S. aureus* | dA9a_s | ~~0.0~~ | ~~0.0~~ | 0.4 | ~~0.0~~ | ~~-0.1~~ |
| Killing rate of *S. aureus* via *Sh* A9 in bacteriocin-resistant *S. aureus* | dA9a_r | ~~0.0~~ | ~~0.0~~ | ~~0.0~~ | ~~0.1~~ | ~~0.0~~ |
| Killing rate of CoNS via ShA9 | dA9h | ~~0.0~~ | ~~0.0~~ | -0.3 | -0.4 | ~~-0.1~~ |
| Inhibitory strength for agr expression of *S. aureus* via *Sh* A9 | bA9s | ~~0.0~~ | ~~0.0~~ | ~~0.0~~ | ~~0.1~~ | ~~0.0~~ |
| Killing rate of *S. aureus* via flucloxacillin | dfa | ~~0.0~~ | ~~0.0~~ | ~~0.0~~ | ~~0.0~~ | 0.3 |
| Killing rate of CoNS via flucloxacillin | dfh | ~~0.0~~ | ~~0.0~~ | ~~0.0~~ | ~~0.0~~ | -0.3 |

**FIGURE S13** Partial rank correlation coefficient (PRCC) between model parameters and %improved EASI by each drug treatment. Open and crossed cells are statistically significant and non-significant PRCC (absolute value >0.1 with adjusted $p$-values <0.05), respectively. Positive PRCC means that virtual patients with a higher value of the parameter achieve a higher %improve EASI by the drug treatment (e.g., $k_3$). Negative PRCC means that virtual patients with a lower value of the parameter achieve a higher %improve EASI by the drug treatment (e.g., $b_2$).

### 6. References


1. George SM, Karanovic S, Harrison DA, et al. Interventions to reduce Staphylococcus aureus in the management of eczema. Cochrane Database Syst Rev. 2019;2019(10):CD003871.
2. Ewing CI, Ashcroft C, Gibbs AC, Jones GA, Connor PJ, David TJ. Flucloxacillin in the treatment of atopic dermatitis. Br J Dermatol. 1998;138(6):1022-1029.
3. Wong SM, Ng TG, Baba R. Efficacy and safety of sodium hypochlorite (bleach) baths in patients with moderate to severe atopic dermatitis in Malaysia. J Dermatol. 2013;40(11):874-880.
4. Leung TH, Zhang LF, Wang J, Ning S, Knox SJ, Kim SK. Topical hypochlorite ameliorates NF-κB-mediated skin diseases in mice. J Clin Invest. 2013;123(12):5361-5370.
5. Sawada Y, Tong Y, Barangi M, et al. Dilute bleach baths used for treatment of atopic dermatitis are not antimicrobial in vitro. J Allergy Clin Immunol. 2019;143(5):1946-1948
6. Gueniche A, Knaudt B, Schuck E, et al. Effects of nonpathogenic gram-negative bacterium Vitreoscilla filiformis lysate on atopic dermatitis: a prospective, randomized, double-blind, placebo-controlled clinical study. Br J Dermatol. 2008;159(6):1357-1363.
7. de Wit J, Totté JEE, van Mierlo MMF, et al. Endolysin treatment against Staphylococcus aureus in adults with atopic dermatitis: A randomized controlled trial. J Allergy Clin Immunol. 2019;144(3):860-863.
8. Myles IA, Earland NJ, Anderson ED, et al. First-in-human topical microbiome transplantation with Roseomonas mucosa for atopic dermatitis. JCI Insight. 2018;3(9):e120608.
9. Nakatsuji T, Gallo RL, Shafiq F, et al. Use of Autologous Bacteriotherapy to Treat Staphylococcus aureus in Patients With Atopic Dermatitis: A Randomized Double-blind Clinical Trial [published online ahead of print, 2021 Jun 16]. JAMA Dermatol. 2021;157(8):978-982.
10. Foelster Holst R, Reitamo S, Yankova R, et al. The novel protease inhibitor SRD441 ointment is not effective in the treatment of adult subjects with atopic dermatitis: results of a randomized, vehicle-controlled study. Allergy. 2010;65(12):1594-1599.
11. Callewaert C, Nakatsuji T, Knight R, et al. IL-4Rα Blockade by Dupilumab Decreases Staphylococcus aureus Colonization and Increases Microbial Diversity in Atopic Dermatitis. J Invest Dermatol. 2020;140(1):191-202.e7.
12. Blauvelt A, de Bruin-Weller M, Gooderham M, et al. Long-term management of moderate-to-severe atopic dermatitis with dupilumab and concomitant topical corticosteroids (LIBERTY AD CHRONOS): a 1-year, randomised, double-blinded, placebo-controlled, phase 3 trial. Lancet. 2017;389:2287-2303
13. Lane P. Handling drop-out in longitudinal clinical trials: a comparison of the LOCF and MMRM approaches. Pharm Stat. 2008;7(2):93-106.
14. Simpson EL, Bieber T, Guttman-Yassky E, et al. Two Phase 3 Trials of Dupilumab versus Placebo in Atopic Dermatitis. N Engl J Med. 2016;375(24):2335-2348.
15. Kabashima K, Matsumura T, Komazaki H, Kawashima M; Nemolizumab-JP01 Study Group. Trial of Nemolizumab and Topical Agents for Atopic Dermatitis with Pruritus. N Engl J Med. 2020;383(2):141-150
16. Simpson EL, Parnes JR, She D, et al. Tezepelumab, an anti-thymic stromal lymphopoietin monoclonal antibody, in the treatment of moderate to severe atopic dermatitis: A randomized phase 2a clinical trial. J Am Acad Dermatol. 2019;80(4):1013-1021
17. Guttman-Yassky E, Pavel AB, Zhou L, et al. GBR 830, an anti-OX40, improves skin gene

signatures and clinical scores in patients with atopic dermatitis. J Allergy Clin Immunol. 2019;144(2):482-493.e7
18. Guttman-Yassky E, Blauvelt A, Eichenfield LF, et al. Efficacy and Safety of Lebrikizumab, a High-Affinity Interleukin 13 Inhibitor, in Adults With Moderate to Severe Atopic Dermatitis: A Phase 2b Randomized Clinical Trial. JAMA Dermatol. 2020;156(4):411-420
19. Silverberg JI, Toth D, Bieber T, et al. Tralokinumab plus topical corticosteroids for the treatment of moderate-to-severe atopic dermatitis: results from the double-blind, randomized, multicentre, placebo-controlled phase III ECZTRA 3 trial. Br J Dermatol. 2021;184(3):450-463.
20. Nakatsuji T, Hata TR, Tong Y, et al. Development of a human skin commensal microbe for bacteriotherapy of atopic dermatitis and use in a phase 1 randomized clinical trial. Nat Med. 2021;27(4):700-709.
21. Wang EB, Shen L, Heathman M, Chan JR. Incorporating Placebo Response in Quantitative Systems Pharmacology Models. CPT Pharmacometrics Syst Pharmacol. 2019;8(6):344-346
22. Miyano T, Irvine AD, Tanaka RJ. A mathematical model to identify optimal combinations of drug targets for dupilumab poor responders in atopic dermatitis. Allergy. 2021;10.1111/all.14870. doi:10.1111/all.14870
23. Koppes SA, Brans R, Ljubojevic Hadzavdic S, Frings-Dresen MH, Rustemeyer T, Kezic S. Stratum Corneum Tape Stripping: Monitoring of Inflammatory Mediators in Atopic Dermatitis Patients Using Topical Therapy. Int Arch Allergy Immunol. 2016;170(3):187-193
24. Williams MR, Costa SK, Zaramela LS, et al. Quorum sensing between bacterial species on the skin protects against epidermal injury in atopic dermatitis. Sci Transl Med. 2019;11(490):eaat8329.
25. Seltmann J, Roesner LM, von Hesler FW, Wittmann M, Werfel T. IL-33 impacts on the skin barrier by downregulating the expression of filaggrin. J Allergy Clin Immunol. 2015;135(6):1659-61.e4
26. Howell MD, Kim BE, Gao P, et al. Cytokine modulation of atopic dermatitis filaggrin skin expression. J Allergy Clin Immunol. 2009;124(3 Suppl 2):R7-R12
27. Syed AK, Reed TJ, Clark KL, Boles BR, Kahlenberg JM. Staphlyococcus aureus phenol-soluble modulins stimulate the release of proinflammatory cytokines from keratinocytes and are required for induction of skin inflammation. Infect Immun. 2015;83(9):3428-3437.
28. Queck SY, Jameson-Lee M, Villaruz AE, et al. RNAIII-independent target gene control by the agr quorum-sensing system: insight into the evolution of virulence regulation in Staphylococcus aureus. Mol Cell. 2008;32(1):150-158.
29. Lambers H, Piessens S, Bloem A, Pronk H, Finkel P. Natural skin surface pH is on average below 5, which is beneficial for its resident flora. Int J Cosmet Sci. 2006;28(5):359-370.
30. Kwaszewska A, Sobiś-Glinkowska M, Szewczyk EM. Cohabitation--relationships of corynebacteria and staphylococci on human skin. Folia Microbiol (Praha). 2014;59(6):495-502.
31. Schröder JM. Antimicrobial peptides in healthy skin and atopic dermatitis. Allergol Int. 2011;60(1):17-24
32. Jack RW, Tagg JR, Ray B. Bacteriocins of gram-positive bacteria. Microbiol Rev. 1995;59(2):171-200.
33. Nakatsuji T, Chen TH, Narala S, et al. Antimicrobials from human skin commensal bacteria protect against Staphylococcus aureus and are deficient in atopic dermatitis.

Sci Transl Med. 2017;9(378):eaah4680.
34. dos Santos Nascimento J, Fagundes PC, de Paiva Brito MA, dos Santos KR, do Carmo de Freire Bastos M. Production of bacteriocins by coagulase-negative staphylococci involved in bovine mastitis. Vet Microbiol. 2005;106(1-2):61-71.
35. Howell MD, Boguniewicz M, Pastore S, et al. Mechanism of HBD-3 deficiency in atopic dermatitis. Clin Immunol. 2006;121(3):332-338
36. Menzies BE, Kenoyer A. Staphylococcus aureus infection of epidermal keratinocytes promotes expression of innate antimicrobial peptides. Infect Immun. 2005;73(8):5241-5244
37. Sieprawska-Lupa M, Mydel P, Krawczyk K, et al. Degradation of human antimicrobial peptide LL-37 by Staphylococcus aureus-derived proteinases. Antimicrob Agents Chemother. 2004;48(12):4673-4679.
38. van Dalen R, De La Cruz Diaz JS, Rumpret M, et al. Langerhans Cells Sense Staphylococcus aureus Wall Teichoic Acid through Langerin To Induce Inflammatory Responses. mBio. 2019;10(3):e00330-19
39. Wanner S, Schade J, Keinhörster D, et al. Wall teichoic acids mediate increased virulence in Staphylococcus aureus. Nat Microbiol. 2017;2:16257.
40. Hanifin JM, Thurston M, Omoto M, Cherill R, Tofte SJ, Graeber M. The eczema area and severity index (EASI): assessment of reliability in atopic dermatitis. EASI Evaluator Group. Exp Dermatol. 2001;10(1):11-18
41. Leung DY. Atopic dermatitis: new insights and opportunities for therapeutic intervention. J Allergy Clin Immunol. 2000;105(5):860-876
42. Grossmann M, Jamieson MJ, Kirch W. Histamine response and local cooling in the human skin: involvement of H1- and H2-receptors. Br J Clin Pharmacol. 1999;48(2):216-222
43. Azimi E, Reddy VB, Lerner EA. Brief communication: MRGPRX2, atopic dermatitis and red man syndrome. Itch (Phila). 2017;2(1):e5.
44. Leung DY, Harbeck R, Bina P, et al. Presence of IgE antibodies to staphylococcal exotoxins on the skin of patients with atopic dermatitis. Evidence for a new group of allergens. J Clin Invest. 1993;92(3):1374-1380.
45. Becker K, Haverkämper G, von Eiff C, Roth R, Peters G. Survey of staphylococcal enterotoxin genes, exfoliative toxin genes, and toxic shock syndrome toxin 1 gene in non-Staphylococcus aureus species. Eur J Clin Microbiol Infect Dis. 2001;20(6):407-409.
46. Yamaguchi M, Sayama K, Yano K, et al. IgE enhances Fc epsilon receptor I expression and IgE-dependent release of histamine and lipid mediators from human umbilical cord blood-derived mast cells: synergistic effect of IL-4 and IgE on human mast cell Fc epsilon receptor I expression and mediator release. J Immunol. 1999;162(9):5455-5465
47. Amin K. The role of mast cells in allergic inflammation. Respir Med. 2012;106(1):9-14
48. Sihto HM, Stephan R, Engl C, Chen J, Johler S. Effect of food-related stress conditions and loss of agr and sigB on seb promoter activity in S. aureus. Food Microbiol. 2017;65:205-212.
49. Bohl T. Lichenification Superimposed on an Underlying Preceding Pruritic Disease. Vulvar Disease. 2019:153-155.
50. Shah DK, Betts AM. Antibody biodistribution coefficients: inferring tissue concentrations of monoclonal antibodies based on the plasma concentrations in several preclinical species and human. MAbs. 2013;5(2):297-305
51. D'Ippolito D, Pisano M. Dupilumab (Dupixent): An Interleukin-4 Receptor Antagonist for Atopic Dermatitis. P T. 2018;43(9):532-535

52. Le Floc'h A, Allinne J, Nagashima K, et al. Dual blockade of IL-4 and IL-13 with dupilumab, an IL-4Rα antibody, is required to broadly inhibit type 2 inflammation. Allergy. 2020;75(5):1188-1204
53. Storn R and Price K. Differential evolution–a simple and efficient heuristic for global optimization over continuous spaces. J Global Optimization. 1997;11(4), 341-359
54. Marino S, Hogue IB, Ray CJ, Kirschner DE. A methodology for performing global uncertainty and sensitivity analysis in systems biology. J Theor Biol. 2008;254:178-196
